# Absorption and Metabolism of Steroidal Alkaloids from Tomato Juice in Healthy Adults: a Pharmacokinetic Study

**DOI:** 10.64898/2026.03.23.26349097

**Authors:** Daniel Do, Maria J. Sholola, Jessica L. Cooperstone

## Abstract

Steroidal alkaloids may be responsible for some of the health benefits of a tomato rich diet, but little is known about their metabolic fate after consumption. The objective of this study was to elucidate the pharmacokinetic parameters of plasma steroidal alkaloids and to define their bioavailability and metabolism following a single tomato containing meal. Healthy subjects (n = 11, 6M/5F) consumed 505 g of tomato juice following a two-week tomato washout and blood plasma were collected post-prandially at 11 time points over 12-hours. Plasma steroidal alkaloids were analyzed using UHPLC-MS. The fractional absorption of steroidal alkaloids was 11.8 ± 7% and over 99% of the absorbed dose were present as metabolized products. The maximum concentration of total plasma steroidal alkaloids in subjects was 406.5 ± 377.0 nmol/L occurring at 6 hours after consumption, with an AUC_0-12hr_ of 2529.0 ± 1644.8 nmol*h/L. Liver S9 enzymatic synthesis of steroidal alkaloid metabolites including trihydroxy-tomatidine and sulfonated dihydroxy-tomatidine improved confidence in compound identification. This study reports the first pharmacokinetic data for tomato steroidal alkaloids, demonstrating moderate absorption and extensive metabolism after tomato juice consumption. These data provide context for future studies investigating the potential role that these compounds may play in human health.

## Introduction

Evidence from epidemiological studies and meta-analyses indicate that increased intake of tomatoes, the second most consumed vegetable in the United States, is inversely associated with the prevalence of and mortality from major chronic diseases such as cardiovascular disease (Cheng et al., 2017; Jacques et al., 2013; Mazidi, Katsiki, et al., 2020) and cancer (Mazidi, Ferns, et al., 2020). Results from clinical trials also suggest beneficial effects of consuming either raw or processed tomato products on markers of cardiovascular disease such as lowered total cholesterol and enhanced high density lipoprotein cholesterol (“good” cholesterol) (Cuevas-Ramos et al., 2013; Silaste et al., 2007; Valderas-Martinez et al., 2016). Even though increased tomato consumption has been found to be beneficial for health, the specific components and the mechanisms that drive bioactivity have yet to be fully elucidated. Historically, the red pigment lycopene has commanded much of this attention. Researchers have found lycopene to be the most effective tomato carotenoid in quenching reactive oxygen species *in vitro* (Di Mascio et al., 1989) and prospective studies show intake to be inversely associated with incidence of prostate cancer (Gann et al., 1999) and acute cardiovascular events (Rissanen et al., 2001). However, lycopene used as a supplement in clinical trials has generally been shown to be less efficacious than those using tomato as whole food (Burton-Freeman & Sesso, 2014). There is a need to study other promising phytochemical candidates to gain a more holistic understanding of what makes tomato consumption healthy. More recently, tomato steroidal alkaloids have emerged as a putative bioactive from tomatoes.

Tomato steroidal alkaloids are a class of cholesterol-derived phytochemicals (Figure 1) with over 100 putative compounds reported (Iijima et al., 2013). These compounds are gaining traction as potential bioactive phytochemicals, especially α-tomatine and tomatidine, for their role in alleviating markers and phenotypes of disease of atherosclerosis (Fujiwara et al., 2007, 2012) and cancer (Lee et al., 2013; Tomsik et al., 2013) in rodent models. The body of *in vitro* research to identify mechanisms of action is also growing (Hsieh et al., 2020; Jeon & Kim, 2019; Kusu et al., 2019; Shi et al., 2013; Shieh et al., 2011; Shih et al., 2009; K. H. Yan et al., 2013). Even so, testing the clinical relevance of steroidal alkaloids is currently premature as we lack a full understanding of their absorption, distribution, metabolism, and excretion (ADME), which are important factors that contribute to efficacy. Some aspects of ADME have been reported in mammals following the consumption of tomato-based interventions. Tomato steroidal alkaloids have been detected in blood and tissue of mice (Cichon et al., 2017; Cooperstone et al., 2017; Dzakovich et al., 2024; Liou et al., 2024), pigs (Sholola et al., 2025), and humans (Hövelmann et al., 2019, 2020) indicating their absorption and in some cases, distribution.

**Figure 1.**
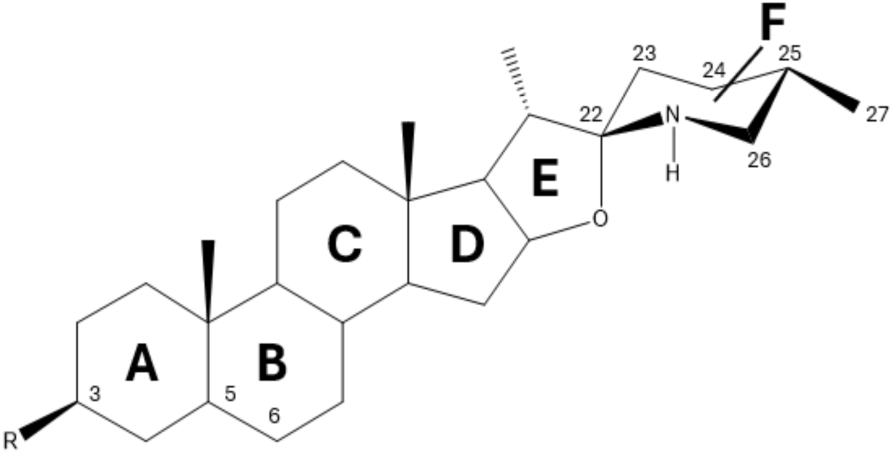
Chemical structure of a tomato steroidal alkaloid backbone. Structural diversity primarily stems from decorations and rearrangements of the F ring. Steroidal alkaloids from food are primarily presented as glycosides with a four sugar (lycotetraose) attachment at the C3 position but when consumed they are primarily found in biological samples in aglycone form.

Steroidal alkaloids found *in vivo* are structurally distinct from what is observed in the diet. Steroidal alkaloids exist primarily as glycosides in food, but nearly all that have been detected *in vivo* have been found as modified aglycones, suggesting either endogenous host or gut microbial metabolism. A study of steroidal alkaloid permeability in a Caco-2 intestinal model showed that aglycones are more permeable than their glycosylated counterparts, and experience extensive metabolism after absorption (Keuter et al., 2024), consistent with previous *in vivo* observations. Additionally, some aspects of excretion have been reported. Steroidal alkaloid metabolites like esculeogenin B sulfonate and hydroxy-esculeogenin B were detectable in urine 48 hours after consumption of 600 g tomato juice (Hövelmann et al., 2020), providing some context to the timeframe of excretion. Two major gaps are still unanswered in humans: what is the bioavailability of tomato steroidal alkaloids from a given dose, and which specific metabolic products are found in circulation? Knowing the amount absorbed from diet and the structural alterations that occur from endogenous metabolism are critical towards our understanding of tomato steroidal alkaloid bioactivity.

To address these gaps, the goal of this study was to determine the fractional absorption of steroidal alkaloids and their pharmacokinetic parameters after consumption of tomato juice in healthy adults. We hypothesized that glycosylated steroidal alkaloids would have moderate oral bioavailability, and that majority of circulating steroidal alkaloids would be metabolized into aglycones prior to undergoing phase I and II metabolism.

## Methods

### Chemicals

Water, acetonitrile, methanol, formic acid (all of liquid chromatography mass spectrometry (LC-MS) grade), phosphoadenosine-5’-phosphosulfate (PAPS, >90% purity), and hydrochloric acid (HCl, technical grade) were purchased from Fisher Scientific (Waltham, MA, United). Trizma^®^ hydrochloride buffer solution, magnesium chloride (MgCl, ≥98% purity), β-nicotinamide adenine dinucleotide 2′-phosphate reduced tetrasodium salt hydrate (NADPH, ≥97% purity), uridine 5′-diphosphoglucuronic acid trisodium salt (UDPGA, >98% purity), acetyl coenzyme A lithium salt (acetyl-CoA, ≥93% purity), ammonium hydroxide (NH_4_OH), tomatidine HCl (≥95% purity), and α-solanine (≥95% purity) were purchased from Sigma-Aldrich (St. Louis, MO, United States). Solanidine (≥99% purity) and α-tomatine (≥90% purity) were purchased from Extrasynthese (Genay, France).

### Study population, design, intervention, and sampling

This pharmacokinetic study is a secondary analysis of samples from a trial that has been published (Cooperstone et al., 2015), and a schematic of the clinical design is detailed in Figure 2. Briefly, 11 subjects (male = 6, female = 5) followed a tomato washout diet for two-weeks prior to their clinical visit. During their visit, subjects consumed tomato juice (505 g) along with a standardized breakfast and blood was taken at 11 time points over 12 hrs: baseline (0 hr) and post-prandially at 0.5, 1, 2, 3, 4, 5, 6, 8, 10 and 12 hrs with a fat-free lunch provided at 4.5 hrs. Blood was spun and plasma was aliquoted and stored at -80 °C until analysis. The clinical study was approved by The Ohio State University Institutional Review Board (IRB, protocol #2012H0189), the Clinical Research Center (Clinical Center for Translational Science ID #1995) and registered on Clinicaltrials.gov (NCT number: NCT01696773).

**Figure 2.**
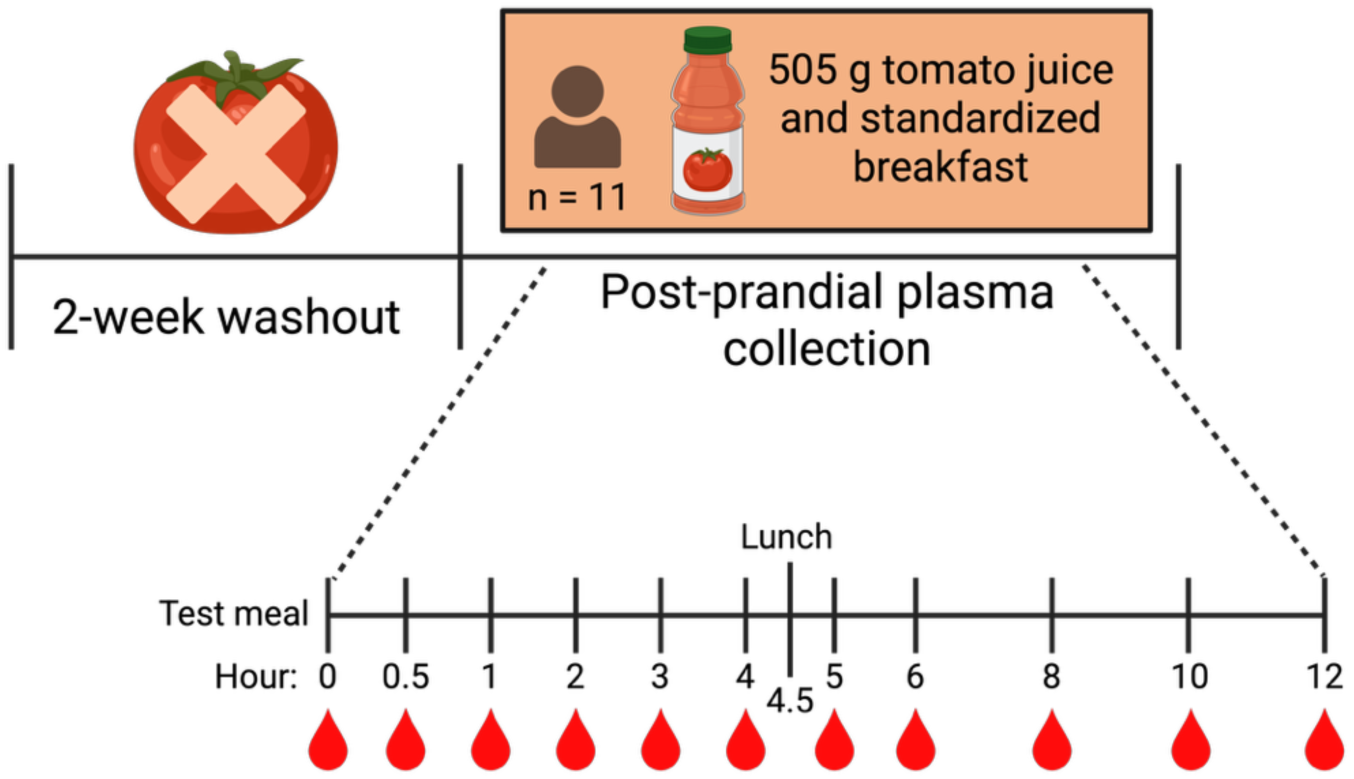
Clinical scheme.

### Quantification of tomato juice steroidal alkaloids using UHPLC-MS/MS

Steroidal alkaloids from the tomato juices were extracted and analyzed using a previously published method (Dzakovich et al., 2020) on a Waters (Milford, MA) Acquity UHPLC interfaced with a Waters (Milford, MA) triple quadrupole detector using multiple reaction monitoring. External standard curves were made using α-tomatine and tomatidine neat solutions with α-solanine and solanidine as internal standards. The linear concentration range was between 5-3821 fmol injected for α-tomatine and 2-1545 fmol injected for tomatidine. Glycosides in the juice for which authentic standards are not available were quantified using the curve of α-tomatine. The ratio of analyte to internal standard signal from sample were referenced to the calibration curve for quantitation.

### Quantification of plasma steroidal alkaloids using UHPLC-QTOF-MS and matrix-matched calibration curves

To accurately reflect the detection response of circulating steroidal alkaloids, matrix-matched standard curves were made using human plasma matrix blanks that contained no background steroidal alkaloids. Plasma samples determined to be devoid of steroidal alkaloids were extracted using a previously detailed method (Sholola et al., 2025). To extract, 100 µL aliquots of plasma were mixed with 500 µL of cold methanol, vortexed, then homogenized for 30 seconds at 1400 RPM using a sample homogenizer (Geno/Grinder 2010, SPEX Sample Prep, Metuchen, NJ, USA), and centrifuged at 21,100 × *g* for 3 min. Five hundred microliters of supernatant were transferred to microfuge tubes, dried in a rotary evaporator (Vacufuge plus, Eppendorf, Hamburg, Germany) and stored at -80 °C until analysis. A stock mixture of α-tomatine and tomatidine standards were prepared at concentrations of 103 pmol/mL and 206 pmol/mL, respectively, and 11 ½ serial dilutions were made from stock. Dried plasma extracts were then reconstituted with 100 µL of 50:50 methanol:water and spiked with 20 µL of the serially diluted tomato steroidal alkaloid standards resulting in 11 in-matrix standards. Each plasma standard mixture was then spiked with 10 µL of α-solanine and solanidine as internal standards at a final concentration of 106 fmol/mL and 561 fmol/mL, respectively. The final concentrations of tomato steroidal alkaloids ranged from 3-3961 fmol/mL for α-tomatine and 7-7923 fmol/mL for tomatidine. Plasma matrix standard solutions were vortexed, transferred to LC vials, and analyzed using UHPLC-QTOF-MS (6545 LC/Q-TOF; Agilent, Santa Clara, CA, USA). A QTOF was used instead of a triple quadrupole instrument to allow for broader discovery of steroidal alkaloid metabolites. Twenty microliter injections were separated using a C18 bridged ethylene hybrid column (2.1 x 100 mm, 1.7 µm; Waters) at 40 °C. The mobile phase gradient with solvent A (water with 0.1% formic acid) and solvent B (acetonitrile with 0.1% formic acid) at a flow rate of 0.4 mL/min was as follows: 95% A for 0.25 min, 95% to 80% A for 1.0 min, 80% to 75% A for 2.5 min, 75% A held for 0.5 min, 75% to 68% A for 1.7 min, 38% to 15% A for 1.7 min, 0% A held for 3.0 min, and a return to 95% A for 2.35 min for a total run time of 13 min. The MS parameters were: gas temperature, 350 °C; gas flow, 10 L/min; nebulizer, 35 psig; sheath gas temperature, 375 °C; sheath gas flow, 11 L/min, and data was collected on full scan in positive ionization mode from 50 to 1700 *m*/*z*. Of the 11 concentrations, six were maintained as calibration points as they were in linear range for both α-tomatine (123-3961 fmol/mL) and tomatidine (247-7923 fmol/mL). Peak areas all were normalized to their internal standard counterpart α-solanine and solanidine, respectively, to correct for any instrument variability.

### Plasma steroidal alkaloid analysis by full scan LC-QTOF-MS and targeted LC-QTOF-MS/MS

Samples (n = 242) were randomized and extracted using the same procedure for creating the blank plasma matrix for standards curves. Once samples were reconstituted to 100 µL, they were spiked with 10 µL of internal standard and 20 µL of additional 50% aqueous methanol for a total volume of 130 µL, to match the solvent volumes used for the standards. Pooled plasma QC samples were also made from a representative pool of samples that included at least one of each subject, collection time, and intervention and were interspersed across the run at fixed intervals to monitor instrument response consistency. Samples were analyzed using the same LC-QTOF-MS parameters detailed above. A list of expected *m*/*z* that correspond to common phase I (dehydrogenation, hydroxylation, acetylation) and phase II (sulfonation, glucuronidation) additions to the parent compounds were curated from available lists of published works (Dzakovich et al., 2024; Sholola et al., 2025) to determine which steroidal alkaloids were potentially present in plasma. Compounds that had an accurate mass match within 10 ppm error of the referenced list, reasonable retention time compared to standard, and were absent in control plasma were putatively assigned as steroidal alkaloid and selected for targeted LC-QTOF-MS/MS. The same chromatography method from above was used for LC-QTOF-MS/MS analysis, and a pre-determined list of masses were fragmented with a collision energy of 40 eV. MS/MS spectra that were consistent with α-tomatine and tomatidine standards or other steroidal alkaloids found in the literature (Hövelmann et al., 2019; Steinert et al., 2020) were kept. All compounds determined to be steroidal alkaloids were quantified using peak area ratios of the analyte to internal standard in reference to the calibration curve.

### In silico assays

There are currently no commercially available standards for the steroidal alkaloid metabolites found in blood. To provide additional confidence to our metabolite identification, steroidal alkaloid metabolites were “bio”-synthesized using a modified procedure from one previously published (Cuparencu et al., 2016). Porcine liver S9 fractions were isolated from pig liver as described (Cuparencu et al., 2016), which served as the source of phase I and II metabolic enzymes. The following cofactors for phase I and II reactions were used: NADPH (oxidation), PAPS (sulfonation), acetyl-CoA (acetylation). To evaluate whether we could create phase I and II steroidal alkaloids metabolites using this system, pure tomatidine standard was used as an aglycone substrate. Briefly, 5 µL of 200 mM tomatidine was mixed with 20 µL S9 fraction (20 mg/mL), 9.40 μL 106 mM MgCl_2_ (source of Mg^2+^ for CYP activity) and 12.5 µL of 40 mM NADPH for phase I reactions and brought up to 500 µL with 0.1 M Tris buffer. For phase II conjugation reactions, either 9.4 µL of 2.5 mg/mL PAPS or 12 µL of 40 mM acetyl CoA was added to the above reaction mixture. All cofactors were prepared in 0.1 M Tris buffer and the final concentration for each is as follows: 1 mM NADPH, 0.5 mM UDPGA, 0.05 mg/mL PAPS, 1 mM acetyl CoA. Other samples were prepared including: a control mixture with no cofactors (i.e., tomatidine, S9 fraction, buffer) to ensure that the cofactors themselves were not contributing to the putatively observed metabolite *m*/*z*, and a control mixture with no substrate (i.e., all cofactors, S9 fraction, and buffer) to ensure that the observed metabolite *m*/*z* was of steroidal alkaloid origin. The mixtures were incubated in a shaking water bath at 37 °C and stirred for 1 hr. The reaction mixture was quenched with cold methanol and centrifuged at 21,100 × *g* for 3 minutes. Aliquots of the supernatant were dried and stored at -80 °C until analysis. Samples were reconstituted with 200 µL of 50% aqueous methanol and were analyzed with UHPLC-QTOF-MS using the same conditions previously described.

The same procedure was then used to investigate phase I and II metabolism of steroidal alkaloids from tomato juices. Steroidal alkaloids extracted from tomato juice were reconstituted in 200 µL 50% aqueous methanol and subjected to acid hydrolysis using a published protocol (Friedman et al., 1998) to cleave the attached sugar moieties to reflect the deglycosylation that occurs *in vivo* (Cichon et al., 2017; Cooperstone et al., 2017; Dzakovich et al., 2024; Hövelmann et al., 2019, 2020; Sholola et al., 2025). The methanolic extracts of tomato juice were incubated in 1 M HCl (4 mL) at 100 °C for 100 min and subsequently cooled. The solution was then neutralized with NH_4_OH, and aliquots were taken for analysis with UHPLC-QTOF-MS to evaluate if hydrolysis was successful. After the hydrolysis was confirmed, the remaining aliquots were subjected to *in silico* reactions above. Targeted LC-QTOF-MS/MS as described above was also performed on the analytes to aid identification.

### Data analysis

Extracted ion chromatograms at the MS1 level were integrated for each metabolite and peak areas were compiled. Data were analyzed using R (version 4.3.3) and RStudio (version 2023.12.1). The pharmacokinetic parameters: C_max_ (max concentration), T_max_ (time of max concentration), AUC_0-12hr_ (area under the curve estimating bioavailability) were calculated using the R package “NonCompart” (Kim et al., 2018) and the function tblNCA. Concentration over time curves were visualized using the package ggplot2 (Wickham, 2016). Fractional absorption was calculated using the following equation:

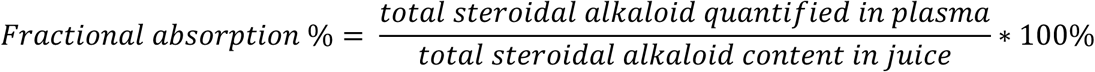

Steroidal alkaloid content in plasma was determined by converting concentration (nmol/L) given by AUC_0-12hr_ to a mass by utilizing the molecular weight of each metabolite and calculating the blood plasma volume for each subject using the Nadler equation (Sharma & Sharma, 2023):

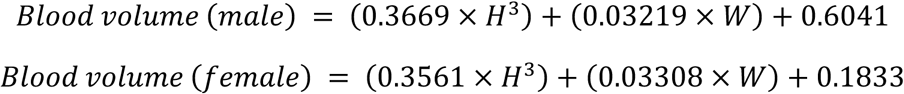

Where *H* is height in meters and *W* is weight in kg. Plasma was assumed to make up 55% of blood volume, therefore, the resulting blood volumes were multiplied by a factor of 0.55 to reflect the samples being analyzed.

## Results

### Nearly all steroidal alkaloids in tomato juice were delivered in glycosylated forms

Six steroidal alkaloids were quantified and the amounts administered in the tomato juice intervention are presented in Table 1. The average total dose of steroidal alkaloids administered to subjects was 2626.8 µg, or ∼2.6 mg. The juice was comprised primarily of α-tomatine and dihydroxy-tomatine, which together constituted 67.3% of the total alkaloids. The next most abundant steroidal alkaloid was hydroxy-tomatine which comprised 21.5% of the total steroidal alkaloid content. Additional steroidal alkaloids detected include dehydro-tomatine and esculeoside B which constituted 0.7-1.2% of the total. Tomatidine, the only aglycone detected in the juice was present at about 0.1% of the total steroidal alkaloid content. Some steroidal alkaloids (e.g., hydroxy-tomatine and dehydro-tomatine) represent the sum of multiple isomeric peaks. The complete list of individual steroidal alkaloid isomers and their concentrations in the dose can be found in Supplemental Table 1. Notably, lycoperosides F, G, and escueloside A, late biosynthetic pathway steroidal alkaloids found in ripe tomatoes (Cárdenas et al., 2015; Dzakovich et al., 2020) were not detected.

**Table 1.** Steroidal alkaloid profile and content (µg) in both low and high dose of tomato juice. Values are represented as means ± SD.

| Steroidal alkaloid | High dose ($\mu\text{g}$) |
| --- | --- |
| α-tomatine | 969.0 ± 116.8 |
| Dihydroxy-tomatine | 798.3 ± 40.0 |
| Hydroxy-tomatine | 564.3 ± 29.7 |
| Dehydro-tomatine | 261.7 ± 22.1 |
| Esculeoside B | 32.5 ± 2.6 |
| Tomatidine | 0.5 ± 0.1 |
| Total | 2626.8 ± 164.5 |

### Steroidal alkaloids and metabolites present in plasma are almost entirely deglycosylated

Steroidal alkaloid metabolites were identified by comparing high resolution mass and retention times to authentic standards and MS/MS fragment pattern matching to those that have been published prior (Cichon et al., 2017; Cooperstone et al., 2017; Dzakovich et al., 2024; Hövelmann et al., 2019, 2020; Sholola et al., 2025). Putative metabolites were also compared to “bio”-synthesized standards (discussed in the next section). A total of 22 individual steroidal alkaloid peaks representing 9 masses (Table 2) were detected in the plasma of subjects 12 hours post-prandially. Of the 6 steroidal alkaloids found in the administered dose, only 2 - α-tomatine and tomatidine - were detected in plasma in the same form as in the juice. The remaining 20 peaks (presenting 7 unique masses) found in plasma were absent in the juice and thus originate from steroidal alkaloid metabolism. Apart from α-tomatine, all annotated metabolites were aglycones and have been previously observed *in vivo* (Cichon et al., 2017; Cooperstone et al., 2017; Dzakovich et al., 2024; Hövelmann et al., 2019, 2020; Liou et al., 2024; Sholola et al., 2025). All metabolites were under 1000 *m/z* which suggest loss of lycotetraose sugar moiety (-618 *m*/*z* from glycoside) and MS/MS corroborate with tomatidine standard for the canonical backbone fragments (273 and 255 *m/z*). Aside from deglycosylation, additional metabolic modifications were postulated based on predicted *m/*z loss from phase I and phase II moieties. These modifications include phase I hydroxylation in mono- (-18 *m/*z), di- (-32 *m/*z), and tri (-50 *m/*z) tomatidine; acetoxylation (-60 *m/z*) in acetoxyhydroxy-tomatidine; and phase II sulfation (-80 *m/z*) in sulfonated -hydroxy-tomatidine, -acetoxyhydroxy-tomatidine, and -dihydroxy-tomatidine and/or esculeogenin B. Multiple peaks were present in the extracted ion chromatogram of several putative masses, all with consistent MS/MS fragmentation. Multiple isomers were observed in blood plasma for hydroxy-tomatidine (2), dihydroxy-tomatidine (up to 3), trihydroxy-tomatidine (up to 6), esculeogenin B (up to 3), hydroxy-esculeogenin B (up to 6), and sulfonated acetoxyhydroxy-tomatidine (2). Isobaric compounds like trihydroxy-tomatidine and hydroxy-esculeogenin B or dihydroxy-tomatidine and esculeogenin B could not be distinguished based on their MS/MS and were grouped indicating this uncertainty.

**Table 2.** Steroidal alkaloids and their metabolites detected in plasma after the consumption of tomato juice, along with their molecular formula, retention time, monoisotopic mass, and characteristic MS/MS fragments. Table is arranged based on elution order. The presence of α-tomatine and tomatidine were confirmed with authentic standards.

| Metabolite | Molecular<br>Formula | Theoretical $m/z$<br>[M + H]<sup>+</sup> | Observed $m/z$<br>[M + H]<sup>+</sup><br>(mass error) | Retention<br>times (min) | MS/MS<br>fragments |
| --- | --- | --- | --- | --- | --- |
| Trihydroxy-tomatidine<br>and/or hydroxy-esculeogenin B | C<sub>27</sub>H<sub>45</sub>NO<sub>5</sub> | 464.3376 | 464.3373<br>(0.6 ppm) | 2.64, 2.90,<br>3.05, 3.12,<br>3.37, 5.05 | 446.3260,<br>428.3138,<br>410.3056,<br>271.2050,<br>253.1948 |
| Sulfonated dihydroxy-tomatidine and/or<br>esculeogenin B | C<sub>29</sub>H<sub>47</sub>NO<sub>8</sub>S | 528.2995 | 528.2985<br>(1.9 ppm) | 3.50, 3.77,<br>4.34, 4.77,<br>5.38 | 510.2871,<br>430.3313,<br>412.3209,<br>394.3099,<br>353.1794,<br>273.2213,<br>255.2110 |
| Dihydroxy-tomatidine<br>and/or esculeogenin B | C<sub>27</sub>H<sub>45</sub>NO<sub>4</sub> | 448.3427 | 448.3428<br>(-0.2 ppm) | 4.17, 4.96,<br>5.50 | 430.3312,<br>412.3214,<br>273.2211,<br>255.2108,<br>161.1323 |
| α-Tomatine | C<sub>50</sub>H<sub>83</sub>NO<sub>21</sub> | 1034.5536 | 1034.5540<br>(-0.4 ppm) | 5.36 | 416.3530,<br>273.2214,<br>255.2102 |
| Sulfonated hydroxy-tomatidine | C<sub>27</sub>H<sub>45</sub>NO<sub>6</sub>S | 512.3045 | 512.3043<br>(0.4 ppm) | 5.45 | 494.3247,<br>414.3385,<br>396.3266,<br>273.1221,<br>255.1109 |
| Sulfonated<br>acetoxhydroxy-<br>tomatidine | C<sub>27</sub>H<sub>45</sub>NO<sub>7</sub>S | 570.3100 | 570.3099<br>(0.2 ppm) | 5.83, 6.78 | 510.2820,<br>472.3413,<br>430.3280,<br>412.3222,<br>353.1771,<br>255.2113 |
| Hydroxy-tomatidine | C<sub>27</sub>H<sub>45</sub>NO<sub>3</sub> | 432.3477 | 432.3477<br>(0.0 ppm) | 5.90, 6.35 | 414.3357,<br>273.2237,<br>161.1264 |
| Acetoxhydroxy-<br>tomatidine | C<sub>29</sub>H<sub>47</sub>NO<sub>5</sub> | 490.3532 | 490.3531<br>(0.2 ppm) | 6.73 | 472.3485,<br>430.3370,<br>412.3214,<br>273.2238,<br>255.2076 |
| Tomatidine | C<sub>27</sub>H<sub>45</sub>NO<sub>2</sub> | 416.3528 | 416.3526<br>(0.5 ppm) | 6.96 | 398.2907,<br>273.2230,<br>255.2126,<br>161.1344 |

### Steroidal alkaloid metabolite identifications were confirmed with *in silico* “bio”-synthesis

Given authentic standards for tomato steroidal alkaloid metabolites are not commercially available, we used pure tomatidine and acid hydrolyzed tomato juice to “bio”-synthesize our own pseudo-standards. To do this, we followed a previously published protocol using a liver S9 metabolic system (Cuparencu et al., 2016) to provide additional confidence in our annotations. We first took the simpler approach of using tomatidine as a starting material to ensure that steroidal alkaloid aglycones could be used as substrate in the liver S9 metabolic system. Tomatidine was incubated with cofactors for phase I (i.e. NADPH) and phase II (i.e. PAPS) reactions separately. One hydroxy-tomatidine isomer (retention time 5.90 min) was synthesized in the phase I incubation, and both the ^13^C isotopic distribution and MS/MS spectra matched what was found in plasma for this specific isomer (Supplemental Figure 1A). Sulfonated tomatidine was synthesized in the phase II incubation, and its MS/MS fragmentation pattern showed characteristic steroidal alkaloid backbone fragments (255 and 273 *m/z*) and loss of sulfur trioxide group (-80 *m/z*) (Supplemental Figure 1B), though this metabolite was not detected in plasma. Once simple phase I and II reaction products could be created in separate reactions from tomatidine, we then co-incubated all cofactors in a single reaction mixture to identify whether more complex metabolites such as sulfonated hydroxy-tomatidine could be synthesized. Co-incubation led to the production of sulfonated hydroxy-tomatidine (Supplemental Figure 1C) with the same retention time, accurate mass, ^13^C isotope distribution, and MS/MS fragments as what was observed in plasma. This metabolic system using tomatidine as substrate were limited to producing only these metabolites (hydroxy-tomatidine and sulfonated tomatidine), while metabolites containing multiple hydroxyl groups were not observed (Supplemental Figures 1D-E).

To better model our *in vivo* system, we used the same style of experiment but substituted the tomatidine with “digested” tomato juice as the substrate. We first hydrolyzed the intervention tomato juice using a published protocol for α-tomatine hydrolysis (Friedman et al., 1998) to create the full range of substrates (i.e., aglycones) that would be available *in vivo*. We found that incubation with 1 N HCl at 100 °C led to nearly complete hydrolysis of all tomato steroidal glycosides to their respective aglycones (Supplemental Figure 2A-F), producing tomatidine, both isomers of hydroxy-tomatidine and 2 of the 3 dihydroxy-tomatidine isomers (retention times 4.96 and 5.50 min) found in plasma. These hydrolysates were subsequently incubated in the liver S9 metabolic system with phase I cofactors, which led to the production of the predominant trihydroxy-tomatidine isomer found in plasma (retention time 2.90 min) (Supplemental Figure 3A). The hydrolysates were then separately incubated with phase II cofactors leading to the production of one isomer of sulfonated hydroxy-tomatidine (retention time 5.45 min) (Supplemental Figure 3B) and one isomer of sulfonated dihydroxy-tomatidine (retention time 5.36 min) (Figure 3C). As a representative example, Figure 3 shows a) extracted ion chromatograms indicating the conversion of dihydroxy-tomatine in juice to dihydroxy-tomatidine by acid hydrolysis along with b) the annotated MS/MS spectra of dihydroxy-tomatidine characterized by iterative water losses (*-*18 *m*/*z*) and the presence of canonical aglycone backbone fragments (273 and 255 *m*/*z*). This newly created dihydroxy-tomatidine was then subsequently used as a substrate to produce sulfonated dihydroxy-tomatidine (Figure 3C) when incubated in the presence of sulfotransferases (in liver S9 fraction) and the cofactor 3’-phosphoadenosine 5’-phosphosulfate. The “bio”-synthesized sulfonated dihydroxy-tomatidine had a mass error within ∼5 ppm of 528.2986 *m/z* and matched the retention time of our putative sulfonated dihydroxy-tomatidine found in plasma (Figure 3C). Figure 3D shows the MS/MS spectra of sulfonated dihydroxy-tomatidine along with the annotation of major fragments including a loss of water (-18 *m*/*z*) and sulfur trioxide (-80 *m/z*) moieties along with fragments consistent with the aglycone backbone (255, 412, and 430 *m*/*z*). No metabolites containing acetoxy groups were observed in the reaction mixtures.

**Figure 3.**
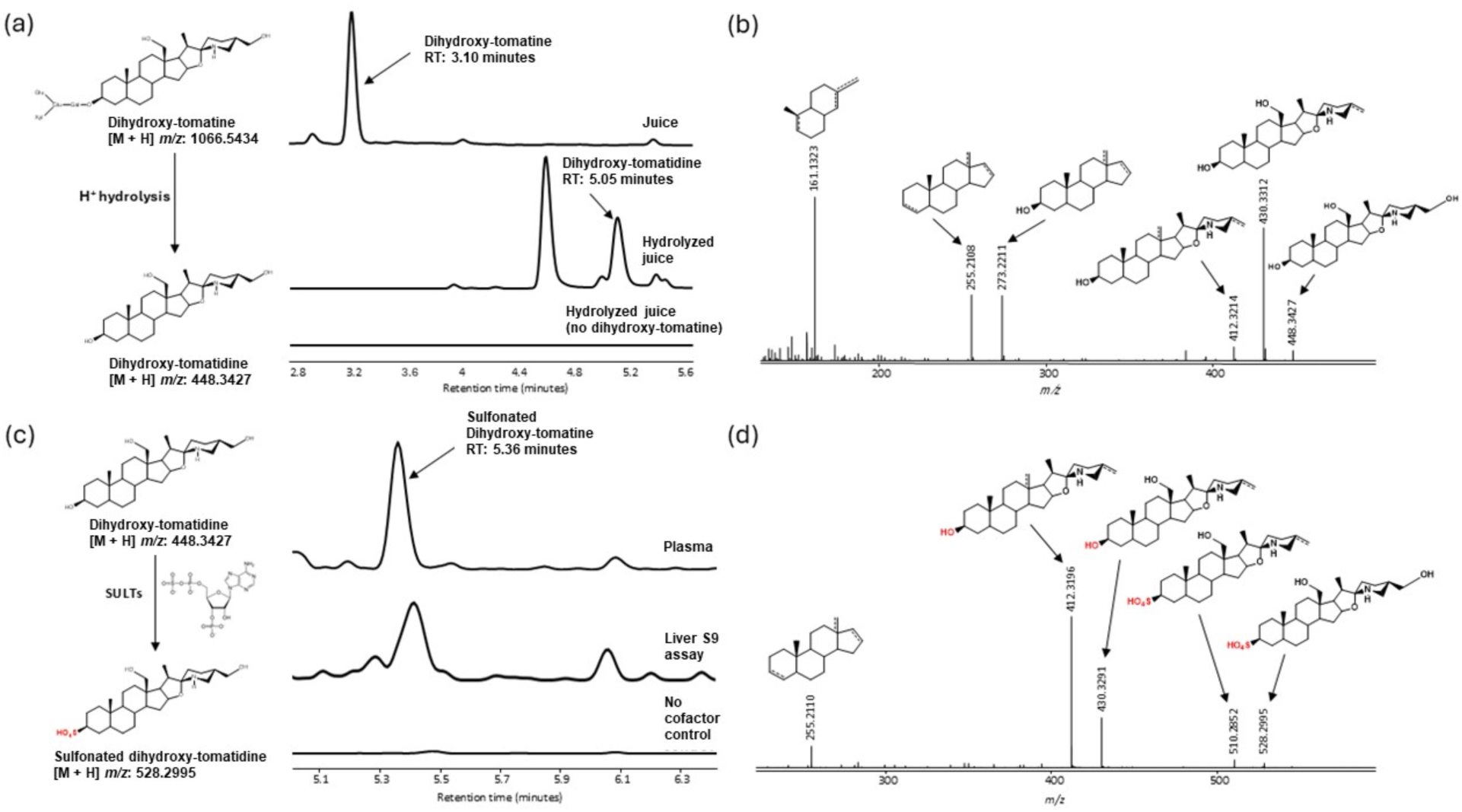
Production of “bio”-synthesized dihydroxy-tomatidine and sulfonated dihydroxy-tomatidine to validate the presence of these compounds in plasma. In detail, a) the conversion of dihydroxy-tomatine from tomato juice to dihydroxy-tomatidine via acid hydrolysis, b) the annotated MS/MS spectra of dihydroxy-tomatidine, c) the conversion of dihydroxy-tomatidine to sulfonated dihydroxy-tomatidine via co-incubation with sulfotransferases contained in the liver S9 fraction and cofactor 3’-phosphoadenosine 5’-phosphosulfate and d) the annotated MS/MS spectra of sulfonated dihydroxy-tomatidine.

### Steroidal alkaloids are moderately absorbed from tomato juice

Fractional absorption here is expressed as the fraction of total steroidal alkaloid metabolites quantified in plasma to the ingested amount from the intervention. Consumption of the tomato juice (∼2 servings of juice containing 2626.8 ± 164.5 μg steroidal alkaloids) led to a fractional absorption (mean ± SD) of 11.8 ± 7.0%, with a range, 2.7 to 22.1% across our 11 subjects.

Total bioavailability is reported as the area under the concentration curve for 12 hours (AUC_0-12hr_), by summing the AUC_0-12hr_ for each metabolite which was 2529.0 ± 1644.8 nmol*h/L. The pharmacokinetic parameters for each steroidal alkaloid metabolite can be found in Table 3. Dihydroxy-tomatidine and/or esculeogenin B were found in plasma in the highest average maximum concentration (C_max_) of 184.6 ± 126.7 nmol/L. Dihydroxy-tomatidine and/or esculeogenin B contributes to around a third of the total bioavailable steroidal alkaloid metabolites, with an AUC_0-12hr_ of 831.9 ± 503.8 nmol/L. Together with the next two highest groups of metabolites in trihydroxy-tomatidine and/or hydroxy-esculeogenin B, and sulfonated dihydroxy-tomatidine and/or esculeogenin B, they contribute to more than 85% of circulating steroidal alkaloids. Sulfonated acetoxyhydroxy-tomatidine contributed ∼10% and the remaining metabolites were more minor. Regarding the time course of the metabolites, most reached their maximum concentration (T_max_) after 6 hours, with α-tomatine having a slightly lower T_max_.

**Table 3.** Pharmacokinetic parameters (AUC_0-12hr_, C_max_, T_max_) of steroidal alkaloid metabolites for both low and high dose. Results are reported as mean ± standard deviation Percent contribution of a metabolite to the total bioavailability is calculated as the AUC_0-12hr_ metabolite/AUC_0-12hr_ total. The table is ordered from lowest to highest abundance metabolites.

| Metabolites | $AUC_{0-12hr}$<br>(nmol*h/L) | % Contribution<br>to total<br>bioavailability | $C_{\max}$ (nmol/L) | $T_{\max}$<br>(h) |
| --- | --- | --- | --- | --- |
| Tomatidine | $2.0 \pm 1.2$ | 0.08% | $0.4 \pm 0.2$ | $7.6 \pm 4.3$ |
| α-Tomatine | $4.2 \pm 4.4$ | 0.17% | $1.2 \pm 2.3$ | $4.5 \pm 2.2$ |
| Hydroxy-tomatidine | $23.1 \pm 17.8$ | 0.91% | $4.6 \pm 3.1$ | $8.7 \pm 2.6$ |
| Sulfonated hydroxy-tomatidine | $26.7 \pm 28.3$ | 1.05% | $6.0 \pm 5.8$ | $10.0 \pm 2.4$ |
| Acetoxyhydroxy-tomatidine | $39.4 \pm 27.2$ | 1.56% | $10.2 \pm 8.9$ | $7.3 \pm 2.1$ |
| Sulfonated acetoxyhydroxy-tomatidine | $259.1 \pm 186.0$ | 10.24% | $52.8 \pm 12.9$ | $9.9 \pm 0.7$ |
| Trihydroxy-tomatidine and/or hydroxy-esculeogenin B | $640.4 \pm 418.6$ | 25.32% | $133.7 \pm 85.9$ | $9.4 \pm 2.4$ |
| Sulfonated dihydroxy-tomatidine and/or esculeogenin B | $694.4 \pm 535.3$ | 27.46% | $149.0 \pm 105.1$ | $9.6 \pm 2.3$ |
| Dihydroxy-tomatidine and/or esculeogenin B | $831.9 \pm 503.8$ | 32.89% | $184.6 \pm 126.7$ | $7.8 \pm 2.6$ |
| Total | $2529.0 \pm 1644.8$ | | | |

The total steroidal alkaloid metabolite plasma concentration over time curve is presented in Figure 4. Each point on the curve is presented as the average (± SEM) total plasma steroidal alkaloid metabolite concentration across subjects at a given post-prandial time. α-Tomatine and tomatidine were detectable in most subjects as early as 30 minutes post-prandially (Supplemental Figure 4A-B). There was also early detection of dihydroxy-tomatidine and/or esculeogenin B, trihydroxy-tomatidine and/or hydroxy-esculeogenin B, and sulfonated acetoxyhydroxy-tomatidine (Supplemental Figure 4G-I) across two subjects at 30 minutes. Average total plasma concentration of steroidal alkaloid metabolites across subjects reached its maximum at 6 hours (Figure 4A) and steroidal alkaloid metabolites were detectable at the end of the collection period. Several metabolites persisted in circulation at relatively high levels at 12 hours: sulfonated dihydroxy-tomatidine at 115.78 ± 78.04 nmol/L (77% of C_max_), followed by trihydroxy-tomatidine and/or hydroxy-esculeogenin B at 83.72 ± 68.22 nmol/L (63% of C_max_) and dihydroxy-tomatidine and/or esculeogenin B at 77.34 ± 57.85 nmol/L (42% of C_max_). Only α-tomatine and tomatidine returned to baseline concentrations (Supplemental Figure 4A-B). High interindividual variability in plasma metabolite concentrations was also observed (Figure 4B). Among subjects there was a 17-fold C_max_ difference between the highest of 1,145.63 nmol/L to the lowest of 67.06 nmol/L, and over a 9 fold difference in AUC_0-12hr_ from 4946.22 nmol*h/L in the highest and 524.49 nmol*h/L in the lowest (Supplemental Table 4).

**Figure 4.**
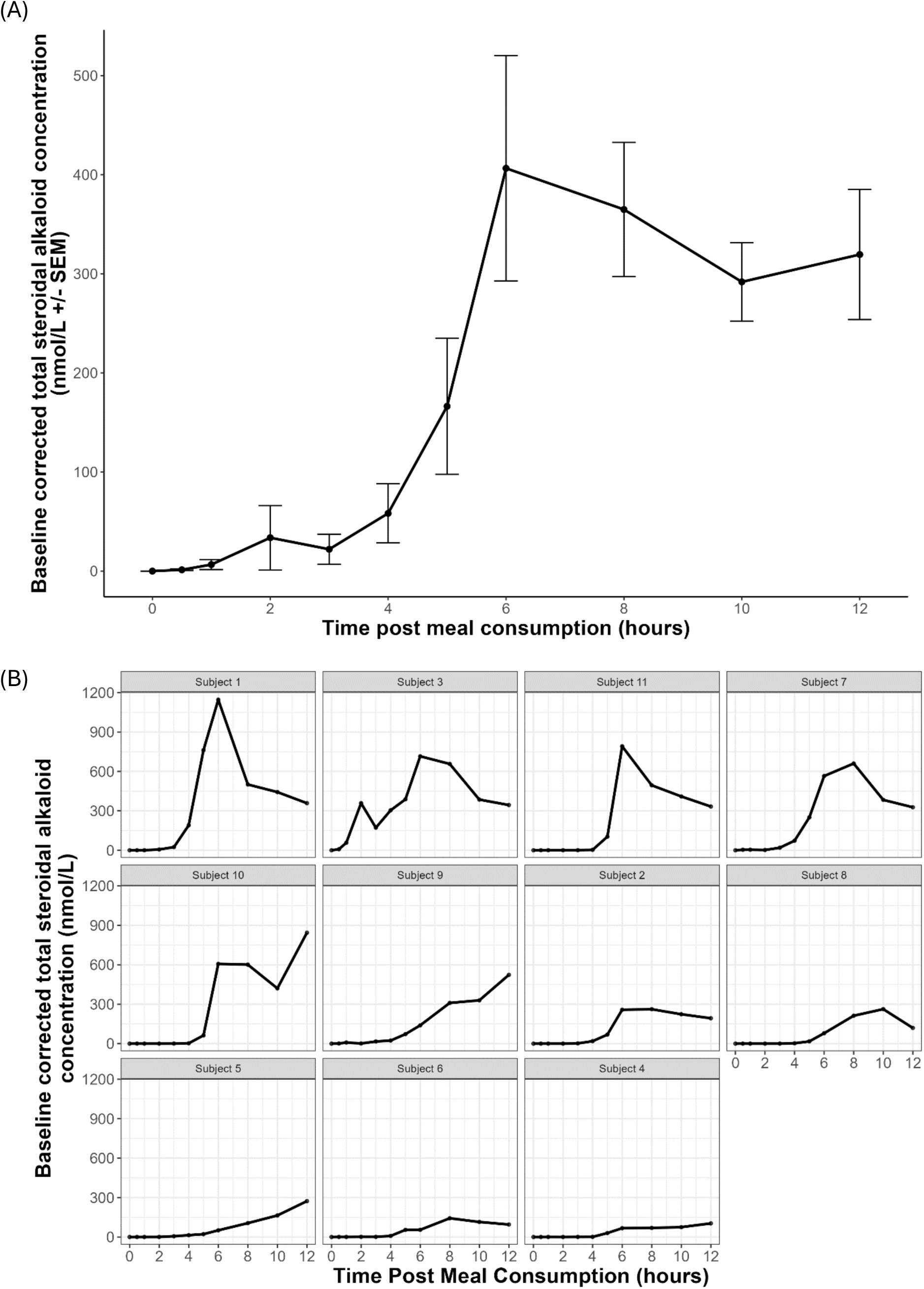
(A) Total plasma steroidal alkaloid concentrations over 12 hour post-prandially after consuming tomato juice intervention. Points depict means while error bars show standard error of the mean (n=11). (B) Individual concentration over time curves for each subject ordered from highest to lowest AUC_0-12hr_.

## Discussion

Here we report for the first time the pharmacokinetics of tomato steroidal alkaloids in humans. Previous reports have posited that tomato steroidal alkaloids were nonabsorbable due to forming insoluble complexes with cholesterol which led to its passing through the gastrointestinal tract and subsequent excretion (Cayen, 1971; Friedman et al., 2000; Wilson et al., 1961). However, recent metabolomics studies have found steroidal alkaloids and their metabolites in different biofluids and tissue across various mammalian species after oral consumption of tomato products, indicating that absorption does occur (Cichon et al. 2017; Cooperstone et al. 2017; Dzakovich et al. 2024; Hövelmann et al. 2019, 2020). These studies though are relative and do not include quantitative alkaloid analysis. Our group recently showed that steroidal alkaloids are present in plasma after short term tomato consumption in pigs (Sholola et al., 2025). In that study, pigs consuming a processing-type tomato (10% diet) accumulated on average 110 nmol/L after two weeks of consumption. Here, we report the quantitation of these established steroidal alkaloid metabolites after 12-hour postprandial consumption of tomato juice, characterizing their pharmacokinetic parameters such as AUC_0-12hr_, C_max_, T_max_ and fractional absorption. “Bio”-synthesized psuedostandards were also created for some of these metabolites which provide greater confidence in their annotation and give insight to their metabolism.

### Circulating steroidal alkaloids are extensively metabolized

For the first time in human plasma, we report the detection and quantification of tomato steroidal alkaloids and their metabolites. We found 22 individual steroidal alkaloid peaks representing 9 unique metabolite masses. The metabolites found in this study are consistent with previous reports in plasma and tissue of mice (Cichon et al., 2017; Cooperstone et al., 2017; Dzakovich et al., 2024; Liou et al., 2024), plasma from pigs (Sholola et al., 2025) and human urine (Hövelmann et al., 2019, 2020), with some minor differences. Steroidal alkaloids from the juice were extensively metabolized prior to reaching systemic circulation. α-Tomatine was the only glycosylated compound detected in plasma and constituted <0.2% of total quantified metabolites despite being the most abundant alkaloid in the juice (37% total alkaloid). α-Tomatine has only been reported *in vivo* in two other cases: pig plasma (Sholola et al., 2025) and mouse serum (Liou et al., 2024). Our lab has previously fed pigs tomato-containing diets for two weeks (Sholola et al., 2025) and is to our knowledge the only other study to both comprehensively profile and quantify circulating steroidal alkaloid metabolites after tomato intervention. In Sholola et al., α-tomatine was present in circulation at a greater proportion (22% total circulating alkaloids) than what we report here (0.2% total plasma alkaloids). Young pigs were used in this study, and the relatively high presence of α-tomatine in plasma was hypothesized to be because of underdeveloped tight junctions in the intestinal epithelium, which enhanced passive diffusion of the glycoside. Healthy adults in general do not have leaky guts, therefore, we would expect to see a lower proportion of α-tomatine in plasma, as demonstrated here. In general, phytochemical glycosides tend to be poorly absorbed and uptake is preceded by deglycosylation, as has been shown with flavonoids (Németh et al., 2003) and saponins (Song et al., 2017). Hydroxylated steroidal alkaloids were the predominant metabolites in circulation which include dihydroxytomatidine/esculeogenin B and its sulfonated counterpart, and trihydroxytomatidine/hydroxyesculeogenin B. Each contributed >25% of total circulated steroidal alkaloids and together these three constituted 85% of all metabolites that were quantified in plasma in this study. This finding is consistent with Sholola et al., where dihydroxytomatidine/esculeogenin B and trihydroxytomatidine/hydroxyesculeogenin B made up the bulk of quantified alkaloids in plasma at 53%, although sulfonated forms were minor (<1%) (Sholola et al., 2025). The lower abundance of sulfonated compounds could be attributed to the lowered activity of phase II sulfation reactions in pig compared to humans (Helke et al., 2016).

Comparing the steroidal alkaloid metabolite profile, we observe the same nine unique masses in human plasma as we do in pigs (Sholola et al., 2025). There were some differences in the isomers observed between the two studies with 22 steroidal alkaloid peaks reported here and 31 in Sholola et al. In our study we found three new isomers of sulfonated dihydroxy-tomatidine (RT: 3.50, 3.77, and 4.34 minutes), while Sholola et al. included additional isomers of hydroxy-tomatidine (2), dihydroxy-tomatidine and/or esculeogenin B (3), trihydroxy-tomatidine and/or hydroxy-esculeogenin B (3), acetoxyhydroxy-tomatidine (2), and sulfonated acetoxyhydroxy-tomatidine (2). Some of the profile differences could be attributed to the type of tomato used in the intervention – heat processed tomato juice here compared to raw, freeze-dried fruits incorporated into the pig diets. This is notable as raw fruits additionally contain lycoperoside F/G/esculeoside A (in this instance, representing 40% of the total alkaloids in the diet) which is absent in our juices. Previous studies have reported that thermal processing of tomatoes, as was the case for our juice, reduced levels of lycoperoside F/G/esculeoside A (Capanoglu et al., 2008; Dzakovich et al., 2020; Katsumata et al., 2011; Steinert et al., 2020). One study demonstrated that heating homogenized cherry tomato to 121 °C for 20 minutes (similar to retort conditions) led to >80% reduction in esculeoside A compared to untreated tomato (Katsumata et al., 2011) while another showed over a two-fold decrease in lycoperoside F/G/esculeoside A in tomato paste compared to fresh fruit (Capanoglu et al., 2008). Acetoxy-tomatine was another alkaloid present in pig plasma but absent here, although this compound has been reported in low levels in processed products (Dzakovich et al., 2020). Despite the absence of acetoxy containing steroidal alkaloids in our juice, metabolites with acetoxy moieties were present in human plasma in this study (∼11% of total alkaloids), indicative of acetylation during metabolism. The difference in intervention length can also influence the plasma steroidal alkaloid profile. Pigs were fed tomatoes for two weeks as opposed to a single dose here, which could elevate the concentration of metabolites that may have been too low to detect from our study. Despite differences in intervention and length between our studies, the prevalence of hydroxylated metabolites in plasma is consistent here between pigs and humans.

### A liver S9 system allows confirmation of steroidal alkaloid metabolite identities and gives insight to their metabolism

Many of the reported *in vivo* steroidal alkaloid metabolites are based on putative identification using relative retention time to α-tomatine and tomatidine standards, accurate mass matching to the predicted monoisotopic mass, and MS/MS fragmentation pattern (Cichon et al., 2017; Cooperstone et al., 2017; Dzakovich et al., 2024; Hövelmann et al., 2019, 2020; Sholola et al., 2025). These identities are by all likelihood correct, though the absence of authentic standards prevents unambiguous identification. In the metabolomics literature, this level of metabolite identification would be considered a level 2 classification, “where at least two orthogonal pieces of information, including evidence that excludes all other candidates” enable a “probable structure” (Blaženović et al., 2018). Without authentic standards, this level 2 classification is the most confident identification possible and often fit for purpose in this area of identifying human metabolites of phytochemicals. Still, there is utility to gain as much confidence as possible in annotations to avoid misidentification leading to misinterpretation, especially when investigating the metabolic fate of phytochemicals of health interest.

Since there are no commercially authentic standards for these steroidal alkaloid metabolites, we used acid hydrolysis along with an established approach to “bio”-synthesize pseudostandards with a liver S9 microsome system (Cuparencu et al., 2016). This brings us closer to a level 1 identification, which we have termed “level 1.5” (Sholola et al., 2026). The liver S9 fraction is efficient because it contains both cytosolic (sulfotransferases, N-acetyl transferases) and microsomal (cytochromes P450’s and uridine 5ʹ-diphospho-glucuronosyltransferase) metabolic enzymes enabling the broad suite of phase I and II reactions, and it is relatively easy to obtain through liver homogenization and centrifugation (Richardson et al., 2016).

The metabolites “bio”-synthesized by the liver S9 system give us insight into how steroidal alkaloid aglycones could be metabolized. We hypothesize that our proposed steroidal alkaloid metabolites could derive from endogenous mammalian metabolism which includes phase I (hydroxylation) and II (sulfation) reactions. Given that deglycosylation is also a major mode of steroidal alkaloid metabolism, it is plausible that this may occur endogenously (via acid hydrolysis in the stomach or enzymatic cleavage via glycosidases in the small intestine) but we are unable to confirm this using the liver S9 system alone. However, more recent work has demonstrated that some of these same metabolites could also be produced by the gut microbiota. Work in a recent preprint has shown that some human colonic microbial strains (including *Alistepes*, *Bacteroides* and *Roseburia*) can perform tomato steroidal alkaloid deglycosylation, hydroxylation, sulfation and acetylation (Liou et al., 2024). Mice fed antibiotics alongside a tomato-containing diet were unable to produce additionally hydroxylated, acetylated, and sulfonated metabolites (Liou et al., 2024). Tomatidine was not detected in serum, cecal contents, or liver and α-tomatine levels were elevated in serum and cecal contents of mice treated with antibiotics – suggesting that in the absence of microbes the metabolism of α-tomatine is severely reduced (Liou et al., 2024). Liou et al. provides strong evidence that this metabolism is microbially mediated but does not preclude additional host metabolism.

We were unable to produce the acetyl-containing metabolites such as acetoxyhydroxy-tomatidine and sulfonated acetoxyhydroxy-tomatidine that were observed *in vivo* with this liver S9 approach. Our findings are consistent with Liou et al. where tomatidine was incubated with liver S9 extracts and acetyl-CoA, and acetylated steroidal alkaloid aglycones were not detected. Acetylated tomatidine was detectable when incubating tomatidine in microbial co-culture of *Roseburia* and *Bacteroides* (Liou et al., 2024) showing the capacity for steroidal alkaloid acetylation by these microbial strains. It is possible that steroidal alkaloids are not modified by endogenous phase II acetylation because the preferred substrates for N-acetyltransferases are aromatic amines (Jancova, 2012).

Our study demonstrates that the liver S9 system is a valid method to “bio”-synthesize steroidal alkaloid metabolites when authentic standards are not commercially available. Our approach of hydrolyzing tomato juice followed by liver S9 incubation provides a comprehensive swath of steroidal alkaloid aglycones that reflect the predominant metabolites found in plasma following tomato consumption. Liou et al. also demonstrates that utilizing microbial co-culture with specific strains of *Roseburia* (deglycosylation) and *Bacteroides* (backbone modification) is another viable way to “bio”-synthesize steroidal alkaloid metabolites. Together, these two systems give insight into how steroidal alkaloids can be metabolized. Hydroxy-tomatidine, dihydroxy-tomatidine, and sulfonated hydroxy-tomatidine can be synthesized by these two systems showing that hydroxylation and sulfonation of steroidal alkaloids can occur by both the host (e.g. CYP enzymes in phase I and SULTs in phase II reactions) and microbes (*Bacteroides* spp.). Trihydroxy-tomatidine and/or hydroxy-esculeogenin B synthesis, which involves the hydroxylation of a dihydroxy-tomatidine and/or esculeogenin B substrate, has only been reported using the liver S9 system but we cannot rule out the possibility of microbial metabolism. Given the current data, it appears that microbes are the only known source for steroidal alkaloid aglycone acetylation.

### Steroidal alkaloids from tomato containing meal are moderately absorbed

In this study, we provided ∼2.6 mg of total steroidal alkaloids administered in 505 g (∼ 0.5 L) of tomato juice. We found that fractional absorption of steroidal alkaloids was on average 11.8% ± 7.0% (range, 2.7 to 22.1%) but may be an underestimation as metabolite concentrations did not return to zero after 12-hours. The average tomato steroidal alkaloid intake can roughly be estimated to be ∼1-4 mg/day based on tomato consumption data (Reimers & Keast, 2016) and steroidal alkaloid content data in a survey of fresh and processed tomato products (Dzakovich et al., 2020). With this context, the juice administered here represents a physiologically relevant dose of steroidal alkaloids that would be achievable with normal tomato consumption.

If we consider oral bioavailability as the fraction of unmodified alkaloid from the juice that reaches systemic circulation, bioavailability for steroidal alkaloids would be nearly zero. As discussed previously, α-tomatine was the only intact glycoside from the juice that could be detected in the plasma. *In vitro* studies using Caco-2 cells found that steroidal alkaloid aglycones are more permeable across the membrane than glycosides, likely because aglycones are more lipophilic and smaller (Keuter et al., 2024). The low plasma concentration of α-tomatine reflects this. Poor oral bioavailability of other structurally related compounds, such as saponins, has been documented. It has been found that 1.2% of ginseng-derived saponins remain intact in mouse plasma after an oral dose of 40 mg/kg (Fu et al., 2022), and similar results were observed in other cases (Hu et al., 2004; Li et al., 2021). The bioavailability is increased when metabolic products of steroidal alkaloids are considered.

Studies over the past decade have shown that steroidal alkaloids are absorbed after consuming tomatoes, but the mechanisms of this absorption are not known. Steroidal alkaloid aglycones share structural similarity to cholesterol so they could also share modes of transport/absorption including the requirement for micellar incorporation for uptake via transporters or passive diffusion. Some studies have shown that co-consumption of tomato steroidal alkaloids with cholesterol leads to reduced uptake of cholesterol suggesting some form of interaction or competition between the two for absorption (Cayen, 1971; Friedman et al., 2000). The breakfast provided with the tomato juice contained negligible amounts of cholesterol, therefore, we do not expect such competition to impact steroidal alkaloid uptake here. As previously stated, steroidal alkaloids should become more permeable across the epithelial intestinal border after their sugar cleavage. Cleavage can occur via endogenous mechanisms including chemical hydrolysis (in the stomach), enzymatic deglycosylation (in the small intestine) and via microbial action (in the colon). Acid hydrolysis experiments have been conducted where α-tomatine was incubated in physiological conditions (1 N HCl at 37 °C) for up to 3 hours (Friedman et al., 1998). No hydrolysis products were found via thin layer chromatography. More powerful and sensitive analytical techniques currently exist for small molecule characterization now, therefore, some contribution of sugar cleavage via gastric digestion cannot fully be ruled out. There is no data on the enzymatic deglycosylation of steroidal alkaloids via endogenous glycosidases in the small intestine to date. However, it has been shown that endogenous β-glucosidases have broad substrate specificity to phytochemical glycosides (Berrin et al., 2002). For saponin glycosides, data suggests that deglycosylation is microbially driven (Dong et al., 2017; Wan et al., 2013; S. Yan et al., 2018). Liou et al. (2024) has shown that *Roseburia* strains can convert α-tomatine to tomatidine as discussed previously. Microbial deglycosylation of steroidal alkaloids has also been supported by a pig cecum model (Kasimir et al., 2023) which found incubation of α-tomatine led to a ∼30% degradation to tomatidine after 240 min. With the current data, we understand that steroidal alkaloid deglycosylation is an important step for their bioavailability given that aglycones constitute nearly all circulating metabolites.

The involvement of microbes in metabolism can also give insight to the high inter-individual variability in fractional absorption of orally consumed steroidal alkaloids. All participants in our study were healthy, non-pregnant, non-smoking, free of malabsorption disorders, and close in age (Supplement Table 2). The intervention was conducted in a highly controlled clinical setting where participants were given the same tomato juice and standardized meal. Therefore, variability in response could be attributed to differences in host (e.g., genetics, microbiome, physiological conditions) (Hasan & Yang, 2019; Kurilshikov et al., 2021) and lifestyle factors (e.g. diet, exercise, sleep) (Conlon & Bird, 2015). Previous work has demonstrated that inter-individual variation in microbiota can impact how α-tomatine is deglycosylated (Liou et al., 2024). α-Tomatine was incubated with 30 different human stool samples and some individuals had little to no capacity for microbial deglycosylation while others show high deglycosylation activity (Liou et al., 2024). Other factors are likely at play, but we can conclude that the composition of the gut microbiota can influence how steroidal alkaloids are absorbed.

Two other aspects of ADME remain, distribution and excretion, which were not captured in this study as we did not collect excreta/tissue samples. As a result, only inferences and speculations regarding these processes can be made based on our findings and ADME information for structurally related compounds. The only reports of steroidal alkaloids in peripheral tissue is in mouse liver (Dzakovich et al., 2024) and skin (Cooperstone et al., 2017). Saponins orally administered to rats can be found in various tissues including stomach, intestine, liver, heart, lung, spleen, and kidney (Chen et al., 2023; Zhang et al., 2025). Excretion of steroidal alkaloids could occur by various modes. Non-absorbed steroidal alkaloids could simply be excreted via feces, while absorbed steroidal alkaloids could be eliminated via urinary and/or biliary excretion. The predominant circulating steroidal alkaloid metabolites were hydroxylated and sulfonated which can derive from either endogenous or microbial metabolism. Regardless of their origin, addition of these moieties enhances polarity and thus enabling renal excretion. This finding is consistent with previous reports of steroidal alkaloids metabolites in urine (Hövelmann et al., 2019, 2020). Additionally, some subjects show the presence of multiple peaks in the metabolite concentration-time curves (Supplemental Figure 4) suggesting the possibility of enterohepatic recirculation. This multi-peak phenomena in the concentration-time curves has also been observed in rats after oral administration of saponins (Zhang et al., 2025). Enterohepatic recirculation involves metabolite incorporation to bile which can be excreted by the liver to the intestine, allowing metabolite re-absorption and prolonged systemic exposure. Biliary excretion can also eliminate metabolites via the feces. Physiochemical properties that allow for biliary excretion include molecular weight of over 400 Da (Varma et al., 2012), a threshold passed by all the observed steroidal alkaloid metabolites.

### A proposed steroidal alkaloid metabolism scheme

A proposed generalized scheme for steroidal alkaloid metabolism has been generated, primarily based on data from this study and others (Liou et al., 2024; Sholola et al., 2025). Once steroidal alkaloids are orally ingested, they encounter acidic conditions in the stomach (Figure 5A). Acid hydrolysis of the lycotetraose sugar moiety of steroidal alkaloids has been demonstrated under high heat conditions well above physiological limits (100 °C) (Mendel Friedman et al., 1998), though we cannot rule out the possibility of partial hydrolysis in physiological conditions. After the stomach, steroidal alkaloids travel to the small intestine where deglycosylation may occur via endogenous glycosidases (Figure 5B). Deglycosylation of steroidal alkaloid glycosides enhances their permeability across the intestinal epithelium (Kasimir et al., 2023), although glycosides are also permeable but much less so (Liou et al., 2024; Sholola et al., 2025). Once the alkaloids pass through the basolateral side of the enterocyte they can travel to the liver via the hepatic portal vein to be further metabolized via phase I and II reactions if necessary (Figure 5D). Some metabolites such as dihydroxy-tomatidine can be derived from the deglycosylation of dihydroxy-tomatine from juice which may not necessitate further metabolism. Any steroidal alkaloids not absorbed in the small intestine pass to the colon where they encounter the gut microbiota (Figure 5C). *Roseburia* strains are capable of deglycosylation while some *Bacteroides* strains can perform various metabolic reactions: hydroxylation, sulfation, and acetylation (Liou et al., 2024). *Bacteroides* are the only known source for steroidal alkaloid aglycone acetylation, but similar metabolites can be obtained via deglycosylation of acetoxy-containing glycosides from diet which include lycoperoside F/G/esculeoside A and acetoxy-tomatine. Gut microbiota-derived metabolites could then be absorbed across the colon epithelium where they can travel to the liver via the hepatic portal vein and may experience additional metabolism prior to reaching systemic exposure (Figure 5D). All remaining steroidal alkaloids not absorbed are presumed to pass through to be excreted via feces.

**Figure 5.**
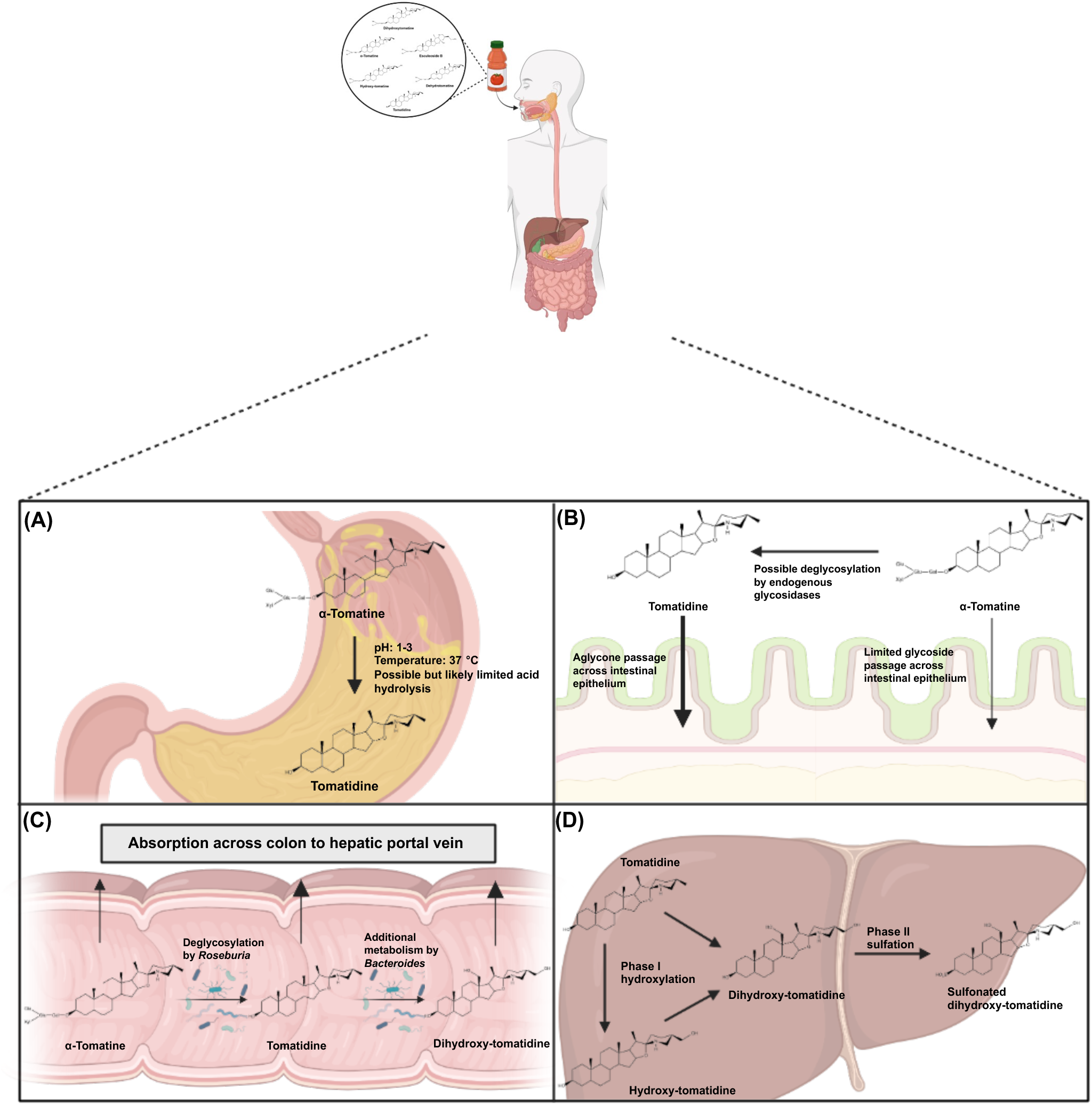
Proposed metabolic scheme of tomato steroidal alkaloids, tracking the plausible metabolic fate of α-tomatine as an example. After oral consumption, steroidal alkaloids are presented in the (A) stomach where acid hydrolysis may occur (α-tomatine to tomatidine conversion here). Once to the (B) small intestine, additional α-tomatine deglycosylation can occur via endogenous glycosidases. Both algycones and glycosides could be absorbed across the intestinal epithelium, but the latter is limited. Absorbed steroidal alkaloids could be transported to the (D) liver via the hepatic portal vein where phase I and II metabolism of aglycones may occur. Steroidal alkaloids not metabolized or absorbed in the small intestine will pass to the (C) colon where these processes can occur via microbial action. In this example, α-tomatine can be converted to tomatidine which can be further metabolized to hydroxylated, sulfated, and acetylated products. Metabolized steroidal alkaloids can be absorbed from the colon for systemic exposure.

## Summary

Here we provide the first report of steroidal alkaloid pharmacokinetics after tomato juice consumption in humans. We have detailed the pharmacokinetic parameters of 9 different steroidal alkaloid masses metabolites, and found that on average, 11.7% of a 2.6 mg dose was absorbed. Absorption also had large interindividual variation, with a range of absorption from 2.7 to 22.1%. Additionally, we “bio”-synthesized 5 steroidal alkaloid metabolites for the first time, providing more conclusive metabolite identification than ever before. Our observation of *in vivo* metabolites corroborate previous studies, and we have established that hydroxyl and sulfate containing metabolites predominate in circulation during a 12 hour post-prandial period. Tomato steroidal alkaloids have been reported to have anti-cancer, anti-inflammatory, and cardioprotective effects and results of this study provide greater context to how they may be affected by our body which could give insight to their bioactivity. Understanding the metabolic fate of tomato steroidal alkaloids brings us closer to elucidating their bioactivity and their role in the health benefits of tomatoes and its phytochemicals.

## Author contributions

Clinical trial operations of this study were executed by J.L.C. as described in previously published work (Cooperstone et al., 2015) and can be referenced on NCT: NCT01696773. Studies were designed by D.D. and J.L.C. Research performed by D.D. and M.J.S. and data analysis was conducted by D.D. and J.L.C. A first draft of the manuscript was written by D.D. and remaining authors provided revisions and feedback. All authors read and approved the final manuscript. J.L.C. has final responsibility for manuscript content.

## Data availability

All relevant data (both averaged and by subject) can be found in the supplemental files of this work.

## Declaration of Generative AI and AI-assisted technologies in the writing process

The author(s) declare that no generative AI or AI-assisted technologies were used in the writing of this manuscript.

## References

Berrin, J. G., McLauchlan, W. R., Needs, P., Williamson, G., Puigserver, A., Kroon, P. A., & Juge, N. (2002). Functional expression of human liver cytosolic β-glucosidase in Pichia pastoris: Insights into its role in the metabolism of dietary glucosides. European Journal of Biochemistry, 269(1), 249–258. 10.1046/j.0014-2956.2001.02641.x

Blaženović, I., Kind, T., Ji, J., & Fiehn, O. (2018). Software tools and approaches for compound identification of LC-MS/MS data in metabolomics. In Metabolites (Vol. 8, Issue 2). MDPI AG. 10.3390/metabo8020031

Burton-Freeman, B. M., & Sesso, H. D. (2014). Whole Food versus Supplement: Comparing the Clinical Evidence of Tomato Intake and Lycopene Supplementation on Cardiovascular Risk Factors. Advances in Nutrition, 5(5), 457. 10.3945/AN.114.005231

Capanoglu, E., Beekwilder, J., Boyacioglu, D., Hall, R., & De Vos, R. (2008). Changes in antioxidant and metabolite profiles during production of tomato paste. Journal of Agricultural and Food Chemistry, 56(3), 964–973. 10.1021/jf072990e

Cárdenas, P. D., Sonawane, P. D., Heinig, U., Bocobza, S. E., Burdman, S., & Aharoni, A. (2015). The bitter side of the nightshades: Genomics drives discovery in Solanaceae steroidal alkaloid metabolism. In Phytochemistry (Vol. 113, pp. 24–32). Elsevier Ltd. 10.1016/j.phytochem.2014.12.010

Cayen, M. N. (1971). Effect of dietary tomatine on cholesterol metabolism in the rat. Journal of Lipid Research, 12(4), 482–490. 10.1016/S0022-2275(20)39498-0

Chen, B., Kuang, G., Wang, Y., Zhang, Y., Wu, Y., Li, Y., Zhang, J., & Zhang, L. (2023). Pharmacokinetic and tissue distribution study of six saponins in the rat after oral administration of Ilex pubescens extract using a validated simultaneous UPLC-qTOF-MS/MS assay. Journal of Pharmaceutical and Biomedical Analysis, 233(April). 10.1016/j.jpba.2023.115431

Cheng, H. M., Koutsidis, G., Lodge, J. K., Ashor, A., Siervo, M., & Lara, J. (2017). Tomato and lycopene supplementation and cardiovascular risk factors: A systematic review and meta-analysis. Atherosclerosis, 257, 100–108. 10.1016/j.atherosclerosis.2017.01.009

Cichon, M. J., Riedl, K. M., Wan, L., Thomas-Ahner, J. M., Francis, D. M., Clinton, S. K., & Schwartz, S. J. (2017). Plasma Metabolomics Reveals Steroidal Alkaloids as Novel Biomarkers of Tomato Intake in Mice. Molecular Nutrition & Food Research, 61(12). 10.1002/MNFR.201700241

Conlon, M. A., & Bird, A. R. (2015). The impact of diet and lifestyle on gut microbiota and human health. Nutrients, 7(1), 17–44. 10.3390/nu7010017

Cooperstone, J. L., Tober, K. L., Riedl, K. M., Teegarden, M. D., Cichon, M. J., Francis, D. M., Schwartz, S. J., & Oberyszyn, T. M. (2017). Tomatoes protect against development of UV-induced keratinocyte carcinoma via metabolomic alterations. Scientific Reports, 7(1), 1–9. 10.1038/s41598-017-05568-7

Cuevas-Ramos, D., Almeda-Valdés, P., Chávez-Manzanera, E., Meza-Arana, C. E., Brito-Córdova, G., Mehta, R., Pérez-Méndez, O., & Gómez-Pérez, F. J. (2013). Effect of tomato consumption on high-density lipoprotein cholesterol level: A randomized, single-blinded, controlled clinical trial. Diabetes, Metabolic Syndrome and Obesity: Targets and Therapy, 6, 263–273. 10.2147/DMSO.S48858

Cuparencu, C. S., Andersen, M. B. S., Gürdeniz, G., Schou, S. S., Mortensen, M. W., Raben, A., Astrup, A., & Dragsted, L. O. (2016). Identification of urinary biomarkers after consumption of sea buckthorn and strawberry, by untargeted LC–MS metabolomics: a meal study in adult men. Metabolomics, 12(2), 1–20. 10.1007/s11306-015-0934-0

Di Mascio, P., Kaiser, S., & Sies, H. (1989). Lycopene as the most efficient biological carotenoid singlet oxygen quencher. Archives of Biochemistry and Biophysics, 274(2), 532–538. 10.1016/0003-9861(89)90467-0

Dong, W. W., Xuan, F. L., Zhong, F. L., Jiang, J., Wu, S., Li, D., & Quan, L. H. (2017). Comparative analysis of the rats’ gut microbiota composition in animals with different ginsenosides metabolizing activity. Journal of Agricultural and Food Chemistry, 65(2), 327–337. 10.1021/acs.jafc.6b04848

Dzakovich, M. P., Goggans, M. L., Thomas-Ahner, J. M., Moran, N. E., Clinton, S. K., Francis, D. M., & Cooperstone, J. L. (2024). Transcriptomics and Metabolomics Reveal Tomato Consumption Alters Hepatic Xenobiotic Metabolism and Induces Steroidal Alkaloid Metabolite Accumulation in Mice. Molecular Nutrition and Food Research, 2300239, 1–11. 10.1002/mnfr.202300239

Dzakovich, M. P., Hartman, J. L., & Cooperstone, J. L. (2020). A High-Throughput Extraction and Analysis Method for Steroidal Glycoalkaloids in Tomato. Frontiers in Plant Science, 11(June), 1–12. 10.3389/fpls.2020.00767

Friedman, M., Fitch, T. E., & Yokoyama, W. E. (2000). Lowering of plasma LDL cholesterol in hamsters by the tomato glycoalkaloid tomatine. Food and Chemical Toxicology, 38(7), 549–553. 10.1016/S0278-6915(00)00050-8

Friedman, M., Kozukue, N., & Harden, L. A. (1998). Preparation and Characterization of Acid Hydrolysis Products of the Tomato Glycoalkaloid α-Tomatine. Journal of Agricultural and Food Chemistry, 46(6), 2096–2101. 10.1021/JF970898K

Fu, X., Chen, K., Li, Z., Fan, H., Xu, B., Liu, M., Guo, L., Xie, Z., Liu, K., Zhang, S., & Kou, L. (2022). Pharmacokinetics and Oral Bioavailability of Panax Notoginseng Saponins Administered to Rats Using a Validated UPLC-MS/MS Method. Journal of Agricultural and Food Chemistry. 10.1021/acs.jafc.2c06312

Fujiwara, Y., Kiyota, N., Hori, M., Matsushita, S., Iijima, Y., Aoki, K., Shibata, D., Takeya, M., Ikeda, T., Nohara, T., & Nagai, R. (2007). Esculeogenin A, a New Tomato Sapogenol, Ameliorates Hyperlipidemia and Atherosclerosis in ApoE-Deficient Mice by Inhibiting ACAT. 10.1161/ATVBAHA.107.147405

Fujiwara, Y., Kiyota, N., Tsurushima, K., Yoshitomi, M., Horlad, H., Ikeda, T., Nohara, T., Takeya, M., & Nagai, R. (2012). Tomatidine, a tomato sapogenol, ameliorates hyperlipidemia and atherosclerosis in ApoE-deficient mice by inhibiting acyl-CoA:cholesterol acyl-transferase (ACAT). Journal of Agricultural and Food Chemistry, 60(10), 2472–2479. 10.1021/jf204197r

Gann, P. H., Ma, J., Giovannucci, E., Willett, W., Sacks, F. M., Hennekens, C. H., & Stampfer, M. J. (1999). Lower prostate cancer risk in men with elevated plasma lycopene levels: Results of a prospective analysis. Cancer Research, 59(6), 1225–1230.

Hasan, N., & Yang, H. (2019). Factors affecting the composition of the gut microbiota, and its modulation. In PeerJ (Vol. 2019, Issue 8). PeerJ Inc. 10.7717/peerj.7502

Helke, K. L., Nelson, K. N., Sargeant, A. M., Jacob, B., McKeag, S., Haruna, J., Vemireddi, V., Greeley, M., Brocksmith, D., Navratil, N., Stricker-Krongrad, A., & Hollinger, C. (2016). Pigs in Toxicology: Breed Differences in Metabolism and Background Findings. Toxicologic Pathology, 44(4), 575–590. 10.1177/0192623316639389

Hövelmann, Y., Jagels, A., Schmid, R., Hübner, F., & Humpf, H. U. (2019). Identification of potential human urinary biomarkers for tomato juice intake by mass spectrometry-based metabolomics. European Journal of Nutrition, 59(2), 685–697. 10.1007/S00394-019-01935-4/FIGURES/6

Hövelmann, Y., Lewin, L., Steinert, K., Hübner, F., & Humpf, H.-U. (2020). Mass Spectrometry-Based Analysis of Urinary Biomarkers for Dietary Tomato Intake. Molecular Nutrition & Food Research, 64, 2000011. 10.1002/mnfr.202000011

Hsieh, M. H., Yang, J. S., Lin, R. C., Hsieh, Y. H., Yang, S. F., Chang, H. R., & Lu, K. H. (2020). Tomatidine Represses Invasion and Migration of Human Osteosarcoma U2OS and HOS Cells by Suppression of Presenilin 1 and c-Raf–MEK–ERK Pathway. Molecules 2020, Vol. 25, Page 326, 25(2), 326. 10.3390/MOLECULES25020326

Hu, J., Reddy, M. B., Hendrich, S., & Murphy, P. A. (2004). Soyasaponin I and sapongenol B have limited absorption by Caco-2 intestinal cells and limited bioavailability in women. Journal of Nutrition, 134(8), 1867–1873. 10.1093/jn/134.8.1867

Iijima, Y., Watanabe, B., Sasaki, R., Takenaka, M., Ono, H., Sakurai, N., Umemoto, N., Suzuki, H., Shibata, D., & Aoki, K. (2013). Steroidal glycoalkaloid profiling and structures of glycoalkaloids in wild tomato fruit. Phytochemistry, 95, 145–157. 10.1016/j.phytochem.2013.07.016

Jacques, P. F., Lyass, A., Massaro, J. M., Vasan, R. S., & D’Agostino, R. B. (2013). Relation of Lycopene Intake and Consumption of Tomato Products to Incident Cardiovascular Disease. The British Journal of Nutrition, 110(3), 545. 10.1017/S0007114512005417

Jancova, P. (2012). Topics on Drug Metabolism. Topics on Drug Metabolism, May. 10.5772/1180

Jeon, S., & Kim, M. M. (2019). Tomatidine inhibits cell invasion through the negative modulation of gelatinase and inactivation of p38 and ERK. Chemico-Biological Interactions, 313, 108826. 10.1016/J.CBI.2019.108826

Kasimir, M., Wolbeck, A., Behrens, M., & Humpf, H. (2023). Intestinal Metabolism of Selected Steroidal Glycoalkaloids in the Pig Cecum Model. ACS Omega, 10.1021/acsomega.3c01990

Katsumata, A., Kimura, M., Saigo, H., Aburaya, K., Nakano, M., Ikeda, T., Fujiwara, Y., & Nagai, R. (2011). Changes in Esculeoside A Content in Different Regions of the Tomato Fruit during Maturation and Heat Processing. J. Agric. Food Chem, 59, 4104–4110. 10.1021/jf104025p

Keuter, L., Wolbeck, A., Kasimir, M., Schürmann, L., & Behrens, M. (2024). Structural Impact of Steroidal Glycoalkaloids : Barrier Integrity, Permeability, Metabolism, and Uptake in Intestinal Cells. Mol. Nutr Food Res., 2300639, 1–14. 10.1002/mnfr.202300639

Kim, H., Han, S., Cho, Y.-S., Yoon, S.-K., & Bae, K.-S. (2018). Development of R packages: “NonCompart” and “ncar” for noncompartmental analysis (NCA). Transl Clin Pharmacol, 26(1), 48–1992. 10.12793/tcp.2018.26.1.xx

Kurilshikov, A., Medina-Gomez, C., Bacigalupe, R., Radjabzadeh, D., Wang, J., Demirkan, A., Le Roy, C. I., Raygoza Garay, J. A., Finnicum, C. T., Liu, X., Zhernakova, D. V., Bonder, M. J., Hansen, T. H., Frost, F., Rühlemann, M. C., Turpin, W., Moon, J. Y., Kim, H. N., Lüll, K., … Zhernakova, A. (2021). Large-scale association analyses identify host factors influencing human gut microbiome composition. Nature Genetics, 53(2), 156–165. 10.1038/s41588-020-00763-1

Kusu, H., Yoshida, H., Kudo, M., Okuyama, M., Harada, N., Tsuji-Naito, K., & Akagawa, M. (2019). Tomatidine Reduces Palmitate-Induced Lipid Accumulation by Activating AMPK via Vitamin D Receptor-Mediated Signaling in Human HepG2 Hepatocytes. Molecular Nutrition & Food Research, 63(22), 1801377. 10.1002/MNFR.201801377

Lee, S. T., Wong, P. F., He, H., Da Hooper, J., & Mustafa, M. R. (2013). Alpha-Tomatine Attenuation of In Vivo Growth of Subcutaneous and Orthotopic Xenograft Tumors of Human Prostate Carcinoma PC-3 Cells Is Accompanied by Inactivation of Nuclear Factor-Kappa B Signaling. PLoS ONE, 8(2).

Li, P., Peng, J., Li, Y., Gong, L., Lv, Y., Liu, H., Zhang, T., Yang, S., Liu, H., Li, J., & Liu, L. (2021). Pharmacokinetics, Bioavailability, Excretion and Metabolism Studies of Akebia Saponin D in Rats: Causes of the Ultra-Low Oral Bioavailability and Metabolic Pathway. Frontiers in Pharmacology, 12. 10.3389/fphar.2021.621003

Liou, C. S., Iakiviak, M., Rajniak, J., Murugkar, P. P., Higginbottom, S. K., Jasper, M., Fischbach, M. A., & Sattely, E. S. (2024). Gut microbiota gate host exposure to metabolites from dietary Solanums. DOI: 10.1101/2024.03.20.584512

Mazidi, M., Ferns, G. A., & Banach, M. (2020). A high consumption of tomato and lycopene is associated with a lower risk of cancer mortality: Results from a multi-ethnic cohort. Public Health Nutrition, 23(9), 1569–1575. 10.1017/S1368980019003227

Mazidi, M., Katsiki, N., George, E. S., & Banach, M. (2020). Tomato and lycopene consumption is inversely associated with total and cause-specific mortality: a population-based cohort study, on behalf of the International Lipid Expert Panel (ILEP). British Journal of Nutrition, 124(12), 1303–1310. 10.1017/S0007114519002150

Németh, K., Plumb, G. W., Berrin, J. G., Juge, N., Jacob, R., Naim, H. Y., Williamson, G., Swallow, D. M., & Kroon, P. A. (2003). Deglycosylation by small intestinal epithelial cell β-glucosidases is a critical step in the absorption and metabolism of dietary flavonoid glycosides in humans. European Journal of Nutrition, 42(1), 29–42. 10.1007/s00394-003-0397-3

Reimers, K. J., & Keast, D. R. (2016). Tomato consumption in the United States and its relationship to the US Department of Agriculture Food Pattern: Results from What We Eat in America 2005-2010. Nutrition Today, 51(4), 198–205. 10.1097/NT.0000000000000152

Richardson, S., Bai, A., A. Kulkarni, A., & F. Moghaddam, M. (2016). Efficiency in Drug Discovery: Liver S9 Fraction Assay As a Screen for Metabolic Stability. Drug Metabolism Letters, 10(2), 83–90. 10.2174/1872312810666160223121836

Rissanen, T. H., Voutilainen, S., Nyyssönen, K., Lakka, T. A., Sivenius, J., Salonen, R., Kaplan, G. A., & Salonen, J. T. (2001). Low serum lycopene concentration is associated with an excess incidence of acute coronary events and stroke: the Kuopio Ischaemic Heart Disease Risk Factor Study. British Journal of Nutrition, 85(6), 749–754. 10.1079/bjn2001357

Sharma, R., & Sharma, S. (2023). Physiology, Blood Volume. StatPearls. https://www.ncbi.nlm.nih.gov/books/NBK526077/

Shi, M. Der, Shih, Y. W., Lee, Y. S., Cheng, Y. F., & Tsai, L. Y. (2013). Suppression of 12-O-Tetradecanoylphorbol-13-Acetate-Induced MCF-7 Breast Adenocarcinoma Cells Invasion/Migration by α-Tomatine Through Activating PKCα/ERK/NF-κB-Dependent MMP-2/MMP-9 Expressions. Cell Biochemistry and Biophysics, 66(1), 161–174. 10.1007/S12013-012-9465-8

Shieh, J. M., Cheng, T. H., Shi, M. Der, Wu, P. F., Chen, Y., Ko, S. C., & Shih, Y. W. (2011). α-Tomatine suppresses invasion and migration of human non-small cell lung cancer NCI-H460 cells through inactivating FAK/PI3K/Akt signaling pathway and reducing binding activity of NF-κB. Cell Biochemistry and Biophysics, 60(3), 297–310. 10.1007/S12013-011-9152-1

Shih, Y. W., Shieh, J. M., Wu, P. F., Lee, Y. C., Chen, Y. Z., & Chiang, T. A. (2009). α-Tomatine inactivates PI3K/Akt and ERK signaling pathways in human lung adenocarcinoma A549 cells: Effect on metastasis. Food and Chemical Toxicology, 47(8), 1985–1995. 10.1016/J.FCT.2009.05.011

Sholola, M. J., Goggans, M. L., Dzakovich, M. P., Francis, D. M., Jacobi, S. K., & Cooperstone, J. L. (2025). Discovery of steroidal alkaloid metabolites and their accumulation in pigs after short-term tomato consumption. Food Chemistry, 463, 141346. 10.1016/j.foodchem.2024.141346

Sholola, M.J., Miller, J., Bilbrey, E.A., Novotny, J.A., Francis, D.A., Mace, T.A., & Cooperstone, J.L. (2026). Tomato-Soy Juice Reduces Inflammation and Modulates the Urinary Metabolome in Adults With Obesity. Molecular Nutrition & Food Research, 70, 70420. 10.1002/mnfr.70420

Silaste, M. L., Alfthan, G., Aro, A., Kesäniemi, Y. A., & Hörkkö, S. (2007). Tomato juice decreases LDL cholesterol levels and increases LDL resistance to oxidation. British Journal of Nutrition, 98(6), 1251–1258. 10.1017/S0007114507787445

Song, P., Zhang, Y., Ma, G., Zhang, Y., Zhou, A., & Xie, J. (2017). Gastrointestinal Absorption and Metabolic Dynamics of Jujuboside A, A Saponin Derived from the Seed of Ziziphus jujuba. Journal of Agricultural and Food Chemistry, 65(38), 8331–8339. 10.1021/acs.jafc.7b02748

Steinert, K., Hövelmann, Y., Hübner, F., & Humpf, H. U. (2020). Identification of Novel Iso-Esculeoside B from Tomato Fruits and LC-MS/MS-Based Food Screening for Major Dietary Steroidal Alkaloids Focused on Esculeosides. Journal of Agricultural and Food Chemistry, 68(49), 14492–14501. 10.1021/ACS.JAFC.0C05906/SUPPL_FILE/JF0C05906_SI_001.PDF

Tomsik, P., Micuda, S., Sucha, L., Cermakova, E., Suba, P., Zivny, P., Mazurova, Y., Knizek, J., Niang, M., & Rezacova, M. (2013). The anticancer activity of alpha-tomatine against mammary adenocarcinoma in mice. Biomed Pap Med Fac Univ Palacky Olomouc Czech Repub, 157(2), 153–161. 10.5507/bp.2013.031

Valderas-Martinez, P., Chiva-Blanch, G., Casas, R., Arranz, S., Martínez-Huélamo, M., Urpi-Sarda, M., Torrado, X., Corella, D., Lamuela-Raventós, R. M., & Estruch, R. (2016). Tomato sauce enriched with olive oil exerts greater effects on cardiovascular disease risk factors than raw tomato and tomato sauce: A randomized trial. Nutrients, 8(3), 1–14. 10.3390/nu8030170

Varma, M. V. S., Chang, G., Lai, Y., Feng, B., El-Kattan, A. F., Litchfield, J., & Goosen, T. C. (2012). Physicochemical property space of hepatobiliary transport and computational models for predicting rat biliary excretion. Drug Metabolism and Disposition, 40(8), 1527–1537. 10.1124/dmd.112.044628

Wan, J. Y., Liu, P., Wang, H. Y., Qi, L. W., Wang, C. Z., Li, P., & Yuan, C. S. (2013). Biotransformation and metabolic profile of American ginseng saponins with human intestinal microflora by liquid chromatography quadrupole time-of-flight mass spectrometry. Journal of Chromatography A, 1286, 83–92. 10.1016/j.chroma.2013.02.053

Wilson, R. H., Poley, G. W., & DeEds, F. (1961). Some pharmacologic and toxicologic properties of tomatine and its derivatives. Toxicology and Applied Pharmacology, 3(1), 39–48. 10.1016/0041-008X(61)90006-0

Yan, K. H., Lee, L. M., Yan, S. H., Huang, H. C., Li, C. C., Lin, H. T., & Chen, P. S. (2013). Tomatidine inhibits invasion of human lung adenocarcinoma cell A549 by reducing matrix metalloproteinases expression. Chemico-Biological Interactions, 203(3), 580–587. 10.1016/j.cbi.2013.03.016

Yan, S., Wei, P. cheng, Chen, Q., Chen, X., Wang, S. cheng, Li, J. ru, & Gao, C. (2018). Functional and structural characterization of a β-glucosidase involved in saponin metabolism from intestinal bacteria. Biochemical and Biophysical Research Communications, 496(4), 1349–1356. 10.1016/j.bbrc.2018.02.018

Zhang, Y., Wang, Y., Qu, B., Liu, L., Yang, J., Hu, X., Chen, Q., Cai, X., & Yan, J. (2025). Pharmacokinetic Profiling and Tissue Distribution of Seven Key Saponins in Rats Following Oral Administration of Panacis Japonici Rhizoma Extracts. ACS Omega, 10(38), 44585–44595. 10.1021/acsomega.5c06902

