## Supplemental Figures for "Absorption and Metabolism of Steroidal Alkaloids from Tomato Juice in Healthy Adults: a Pharmacokinetic Study"

**Supplementary Figures**

**Supplemental Figure 1.** To validate if steroidal alkaloid metabolites could be “bio-”synthesized using a liver S9 microsome metabolic system tomatidine standard was first used as substrate. **The synthesis of steroidal alkaloid metabolites were confirmed with retention time matching with metabolites found in plasma, mass fragmentation, and mass accuracy**. a) We first show that tomatidine can undergo phase I hydroxylation to produce hydroxytomatidine, b) and also phase II sulfation to produce sulfonated tomatidine. c) When tomatidine was incubated with both phase I and II cofactors, sulfonated hydroxytomatidine was produced. Steroidal alkaloid metabolites containing multiple hydroxyl groups are found in highest concentration in plasma, but neither d) dihydroxytomatidine nor e) trihydroxytomatidine could be synthesized from tomatidine when incubated with phase I cofactors.

Retention time (minutes)

5.4

5.6

5.8

6

6.2

6.4

6.6

6.8

7

**Tomatidine**

**standard**

**No cofactor**

**control**

**Liver S9**

**assay**

496.3107

497.3128

498.3127

499.3102

*m/z*

496

496.4

496.8

497.2

497.6

498

498.4

498.8

499.2

499.6

*m/z*

240

280

320

360

400

440

480

520

255.2104

273.2215

353.1780

398.3415

416.3520

496.3092

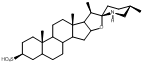

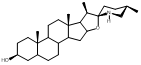

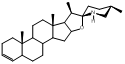

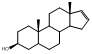

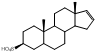

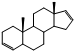

432.3473

*m/z*

432

432.5

433

433.5

434

434.5

433.3502

434.3529

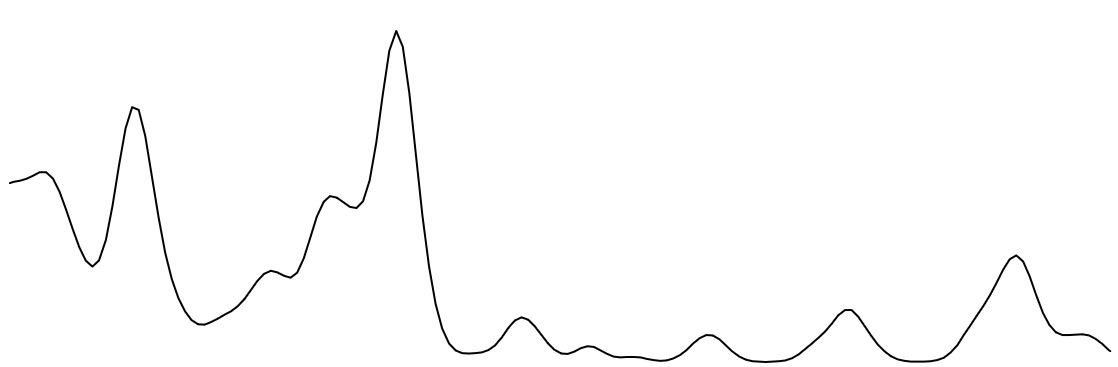

Retention time (minutes)

5.6

5.8

6

6.2

6.4

6.6

6.8

**No cofactor**

**control**

**Tomatidine**

**standard**

**Liver S9**

**assay**

**Plasma**

**Hydroxytomatidine**

432.3467

271.2051

414.3356

253.1953

*m/z*

240

260

280

300

320

340

360

380

400

420

440

460

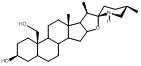

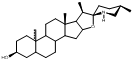

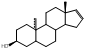

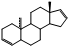

(B)

(A)

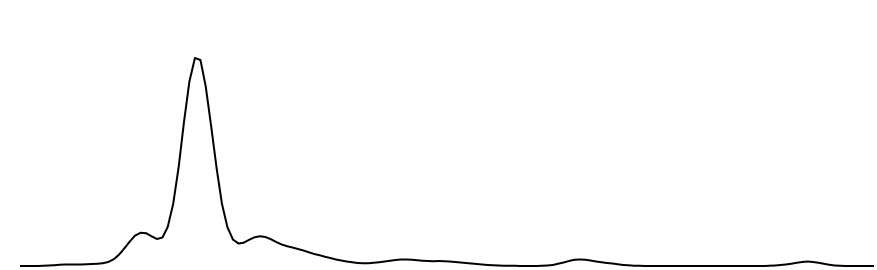

512.3037

*m/z*

512

512.5

513

513.5

514

514.5

515

513.3059

514.2985

515.3079

Retention time (minutes)

5.1

5.3

5.5

5.7

5.9

6.1

6.3

**No cofactor**

**control**

**Tomatidine**

**standard**

**Liver S9**

**assay**

**Plasma**

**Sulfonated**

**hydroxytomatidine**

253.1945

*m/z*

240

260

280

300

320

340

360

380

400

420

440

460

480

500

520

351.1642

396.3259

414.3359

432.3460

512.3030

494.2883

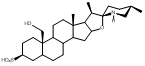

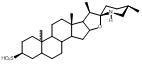

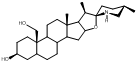

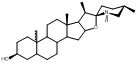

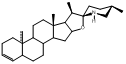

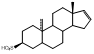

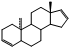

Retention time (min)

4.9

5

5.1

5.2

5.3

5.4

5.5

5.6

5.7

5.8

4.8

**Liver S9**

**assay**

**No cofactor**

**control**

**Plasma**

Retention time (min)

2.5

2.7

2.9

3.1

3.3

3.5

3.7

3.9

4.1

4.3

4.5

**Liver S9**

**assay**

**No cofactor**

**control**

**Plasma**

**Tomatidine standard**

**Tomatidine standard**

**Dihydroxytomatidine**

**Trihydroxytomatidine**

(D)

(E)

(C)

**Supplemental Figure 2.** Tomato steroidal alkaloids in juice were acid hydrolyzed to produce aglycones that are present for *in vivo* metabolism. Deglycosylation of glycoside was confirmed by comparing the extracted ion chromatogram for the glycoside and its respective aglycone in juice and hydrolyzed juice samples. All glycosides in juice were found to be hydrolyzed upon incubation with acid validated with chromatography and MS/MS fragmentation for: α-tomatine to tomatidine conversion (a-b), hydroxytomatine to hydroxytomatidine conversion (c-d), dihydroxytomatine to dihydroxytomatidine conversion (e-f)

(A)

(B)

(C)

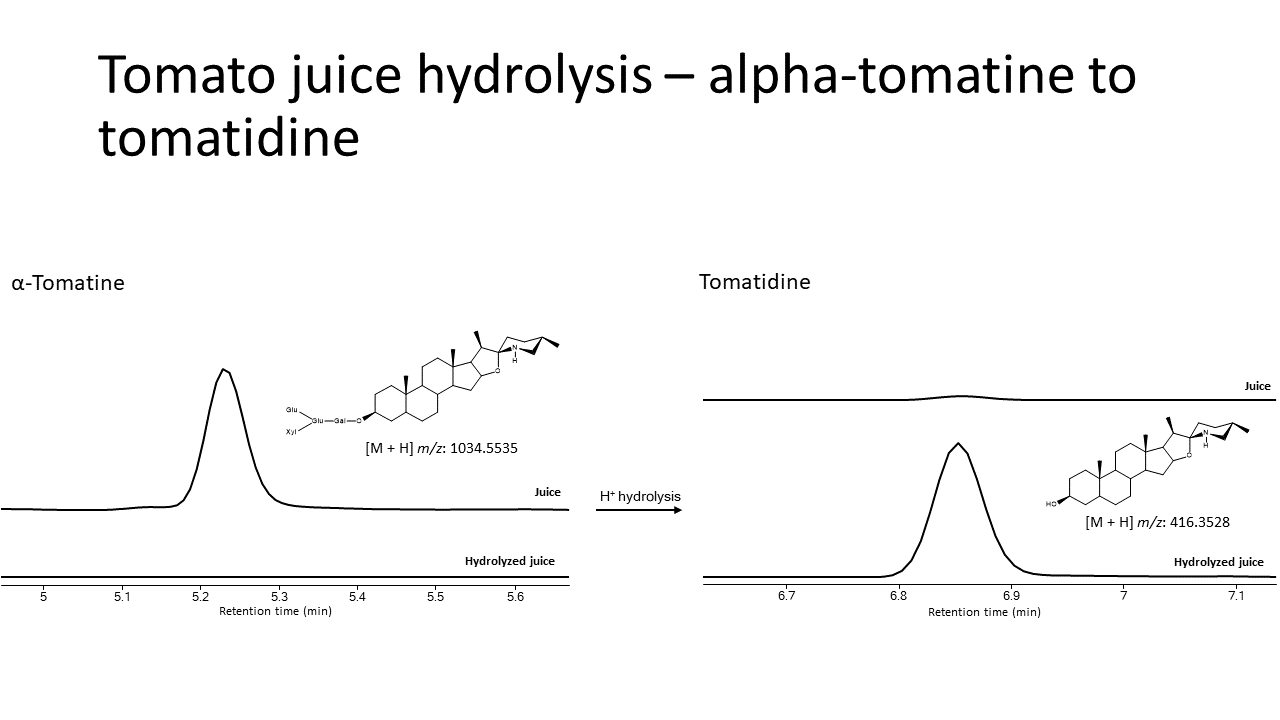

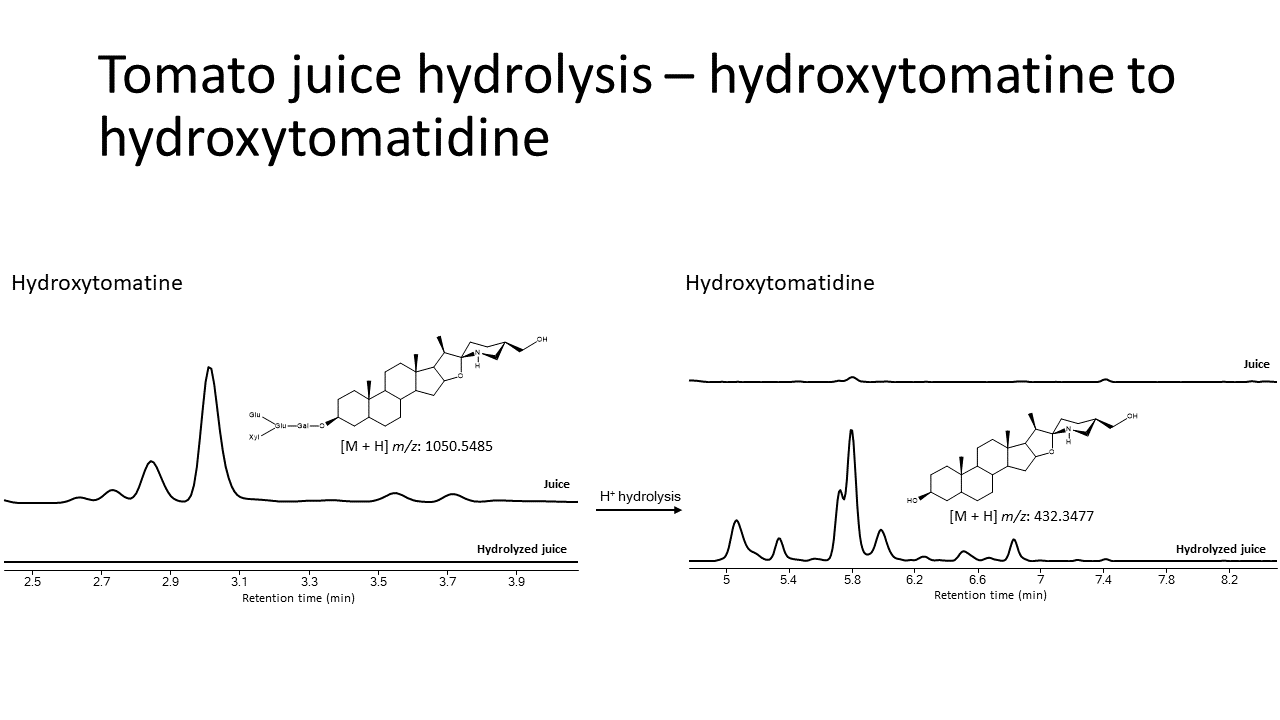

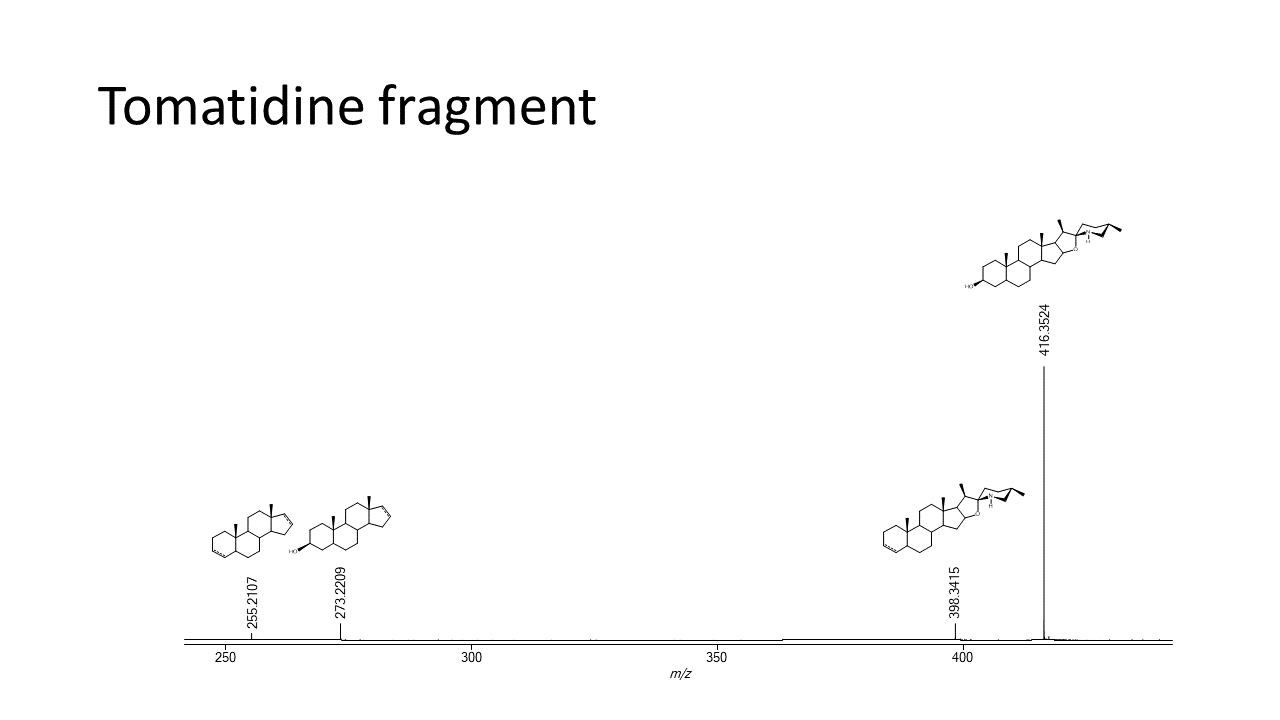

(D)

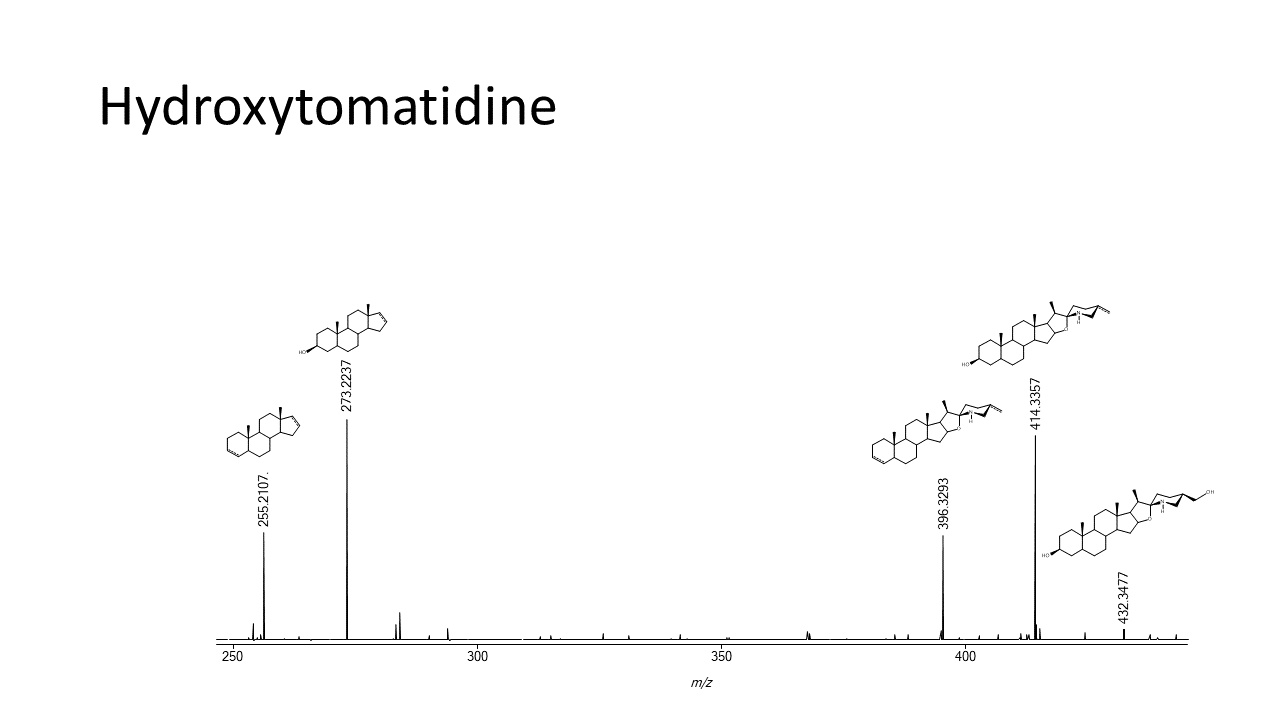

(E)

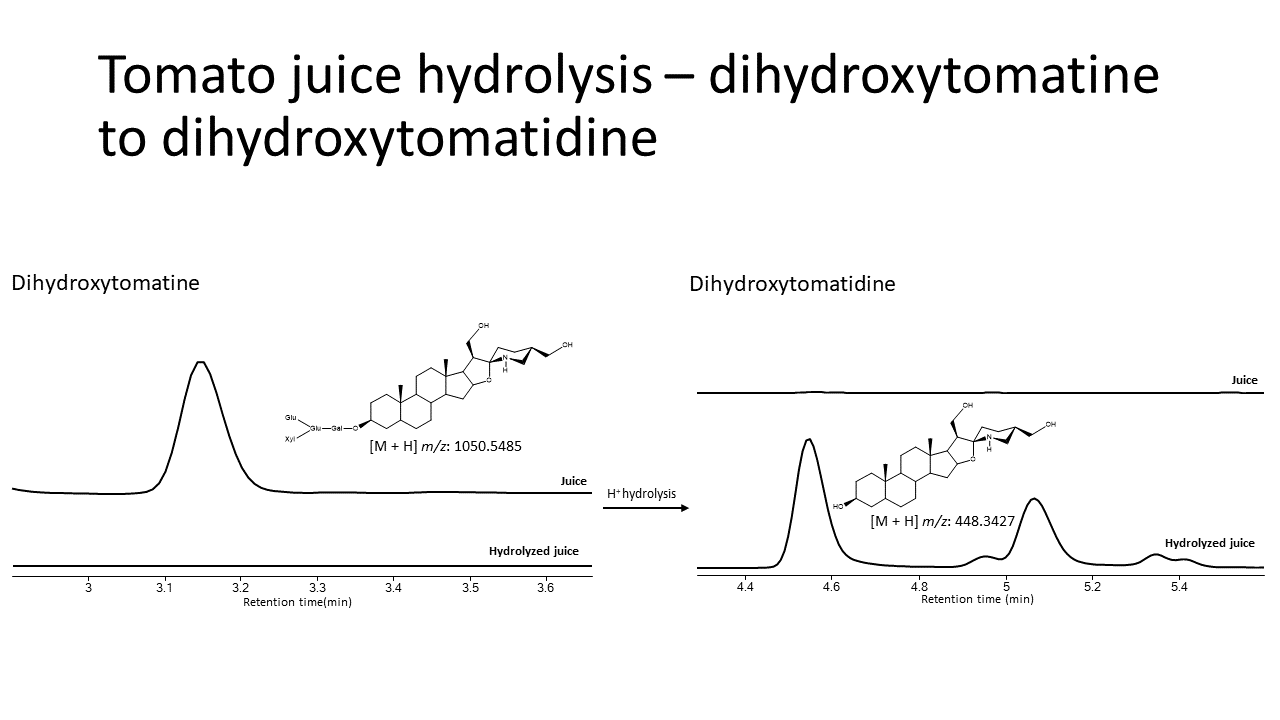

(F)

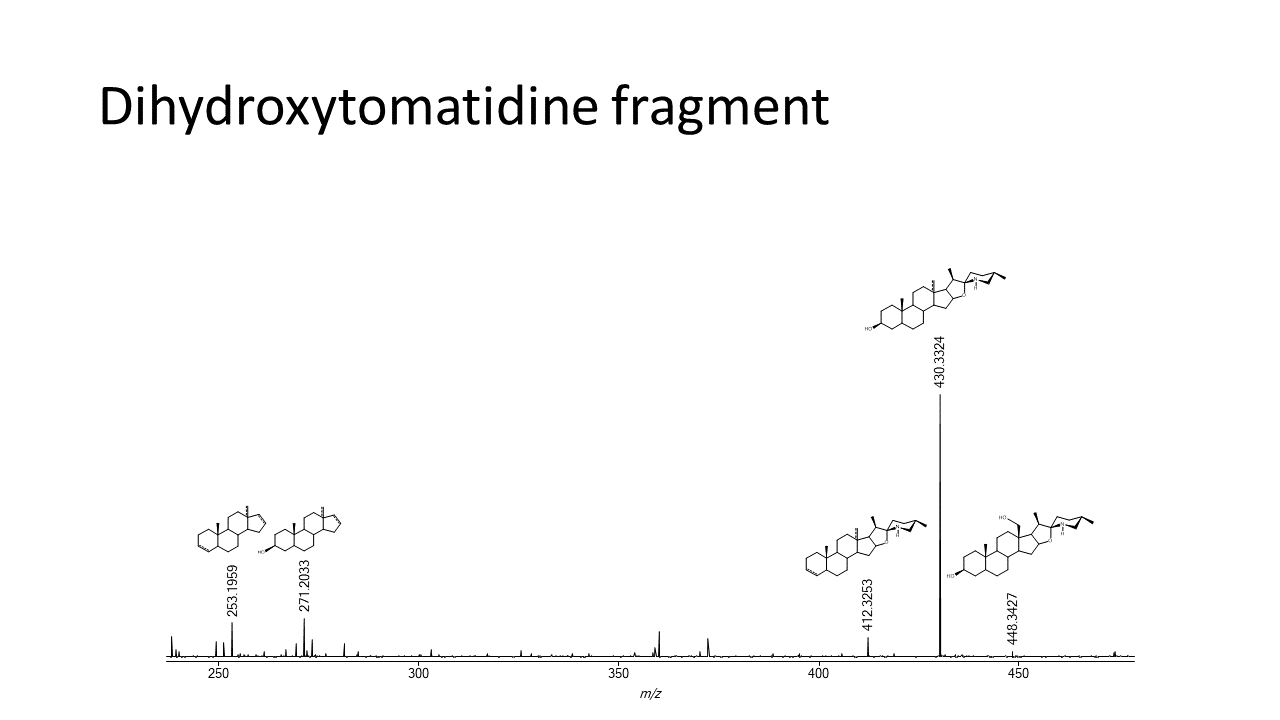

**Supplementary Figure X**. Total steroidal alkaloid concentration over time curves for each subject (a) high dose, (b) low dose.

**Supplemental Figure 3.** The tomato steroidal alkaloid aglycones produced by acid hydrolysis were incubated in the liver S9 system to recapitulate phase I and phase II metabolites not observed in hydrolysis alone. Two metabolites were “bio”-synthesized from the hydrolyzed aglycone substrates, a) trihydroxytomatidine presumably by the hydroxylation of dihydroxytomatidine, and b) sulfonated hydroxytomatidine by the sulfation of hydroxytomatidine. The retention time of the “bio”-synthesized metabolites align with what is observed in plasma for the extracted ion chromatogram for the respective *m/z*, and the compounds produced characteristic mass fragmentation patterns expected for steroidal alkaloid metabolites.

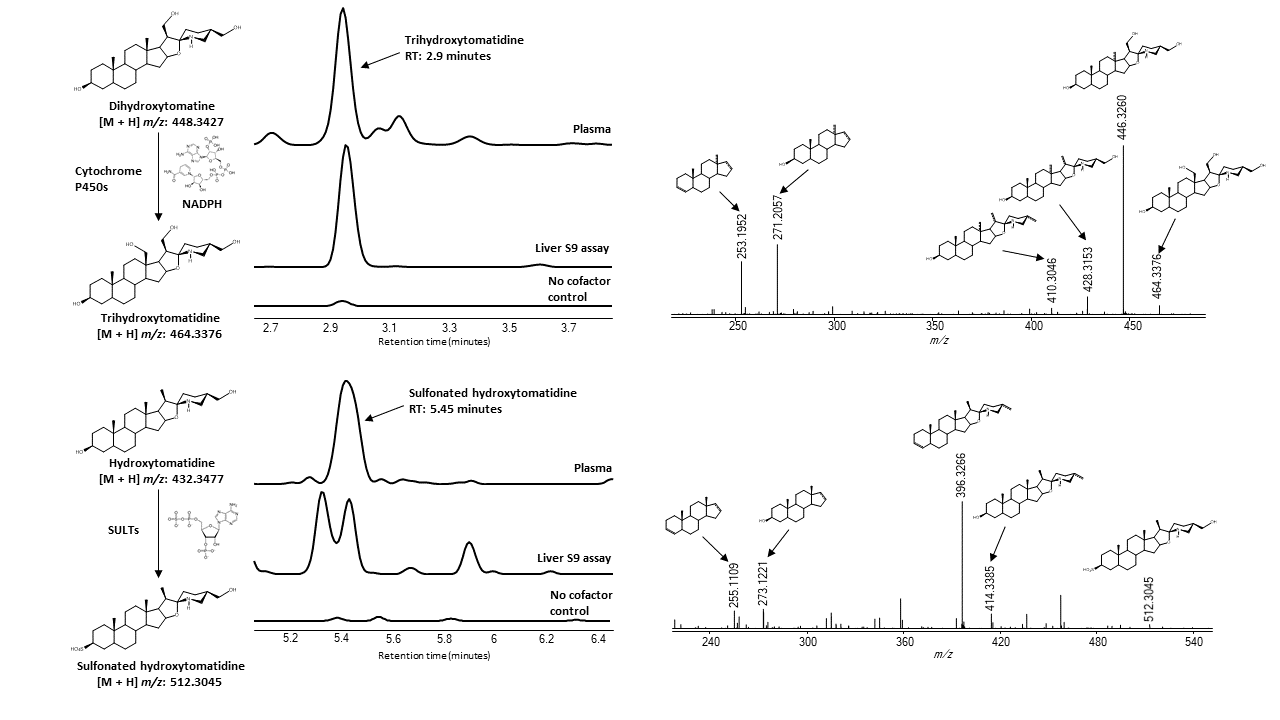

(B)

(A)

**Supplemental Figure 4.** Subject concentration over time curves for each metabolite (A-I). Metabolites with isomers were consolidated into aggregate concentration.

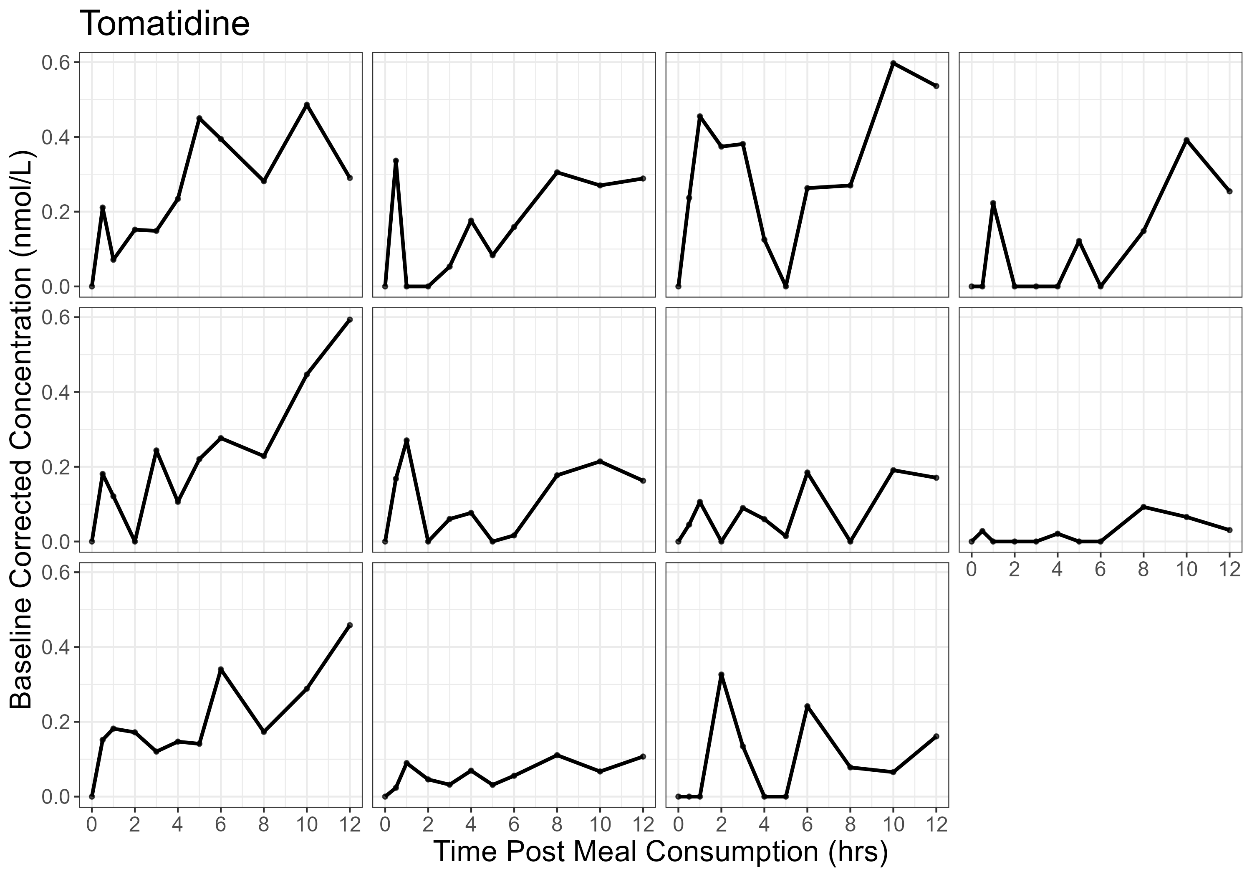

(A)

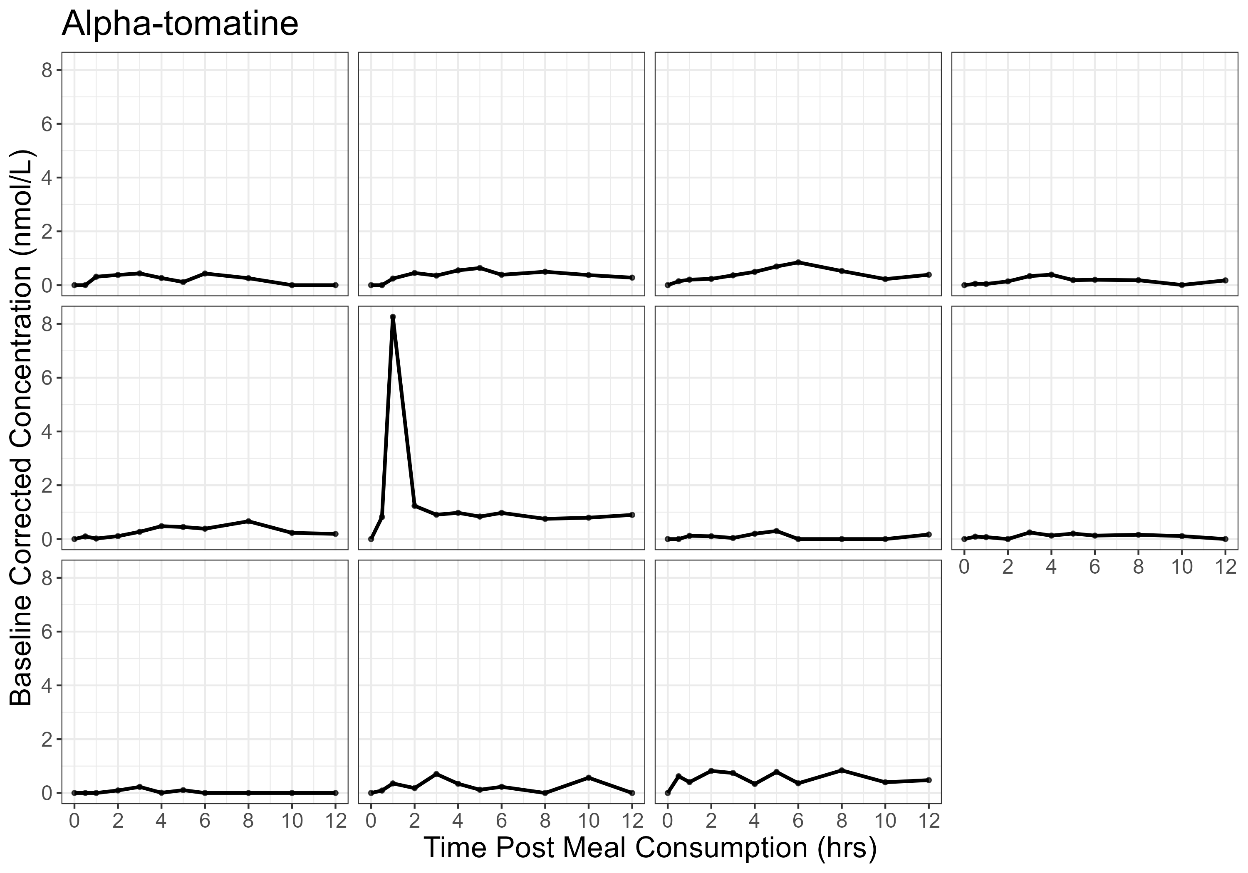

(B)

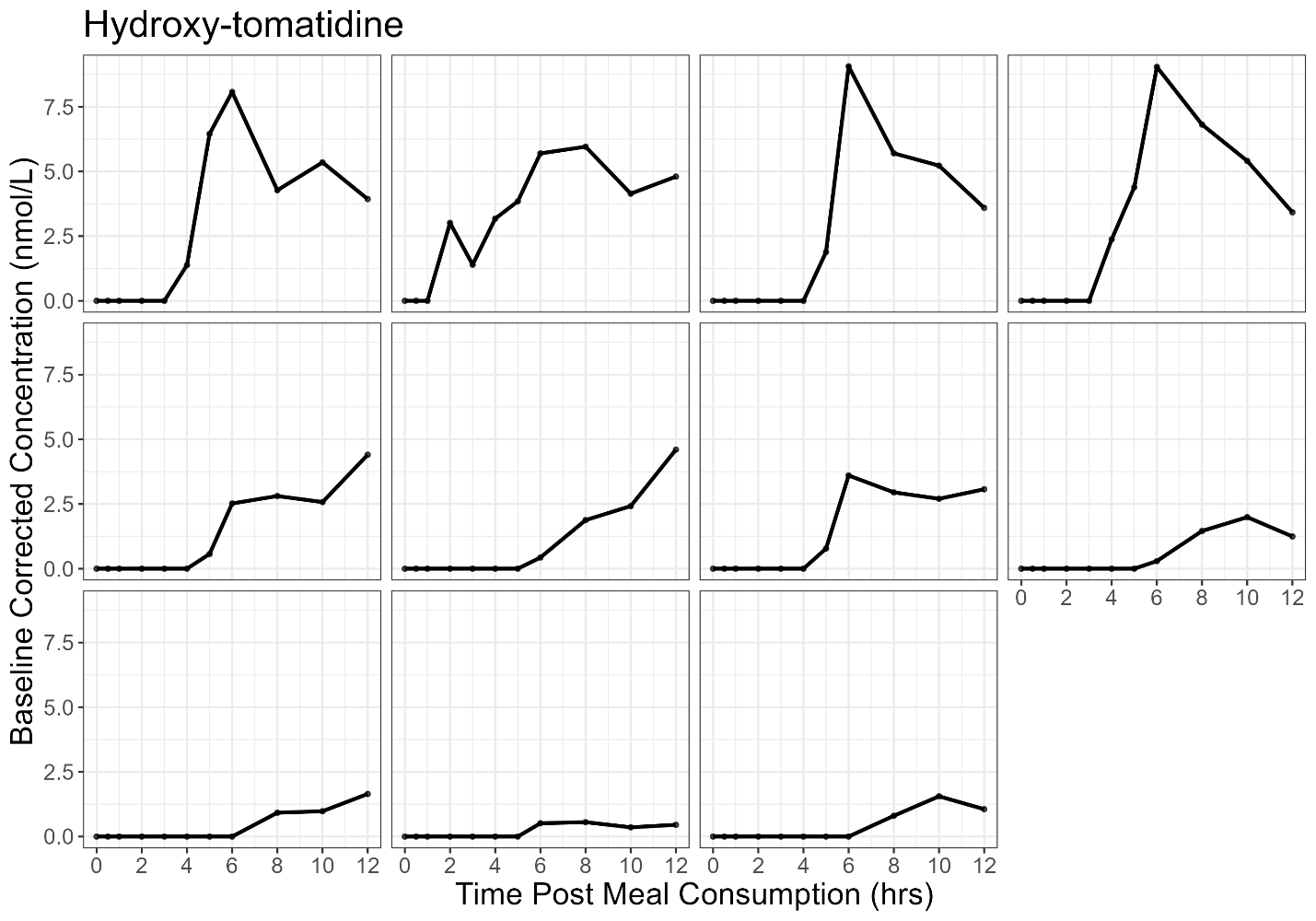

(C)

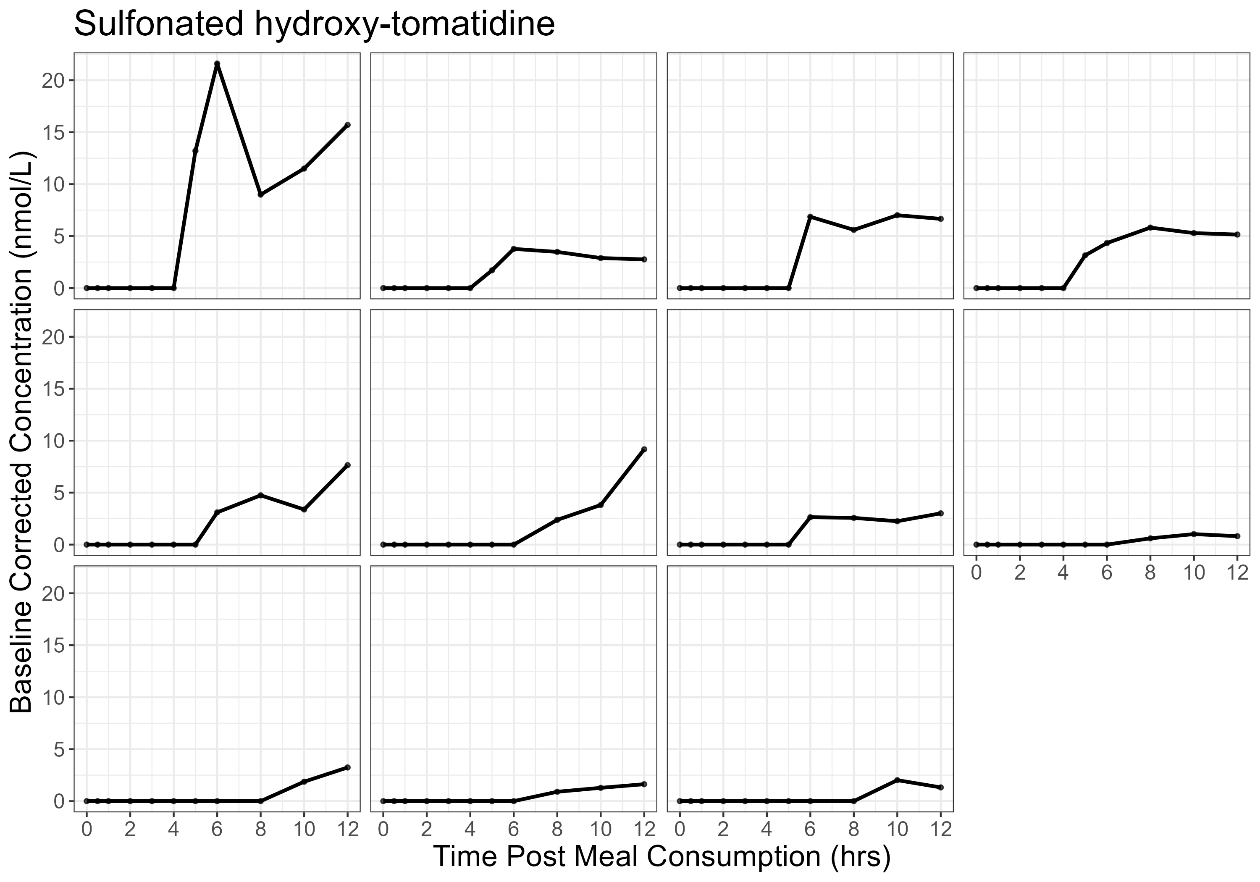

(D)

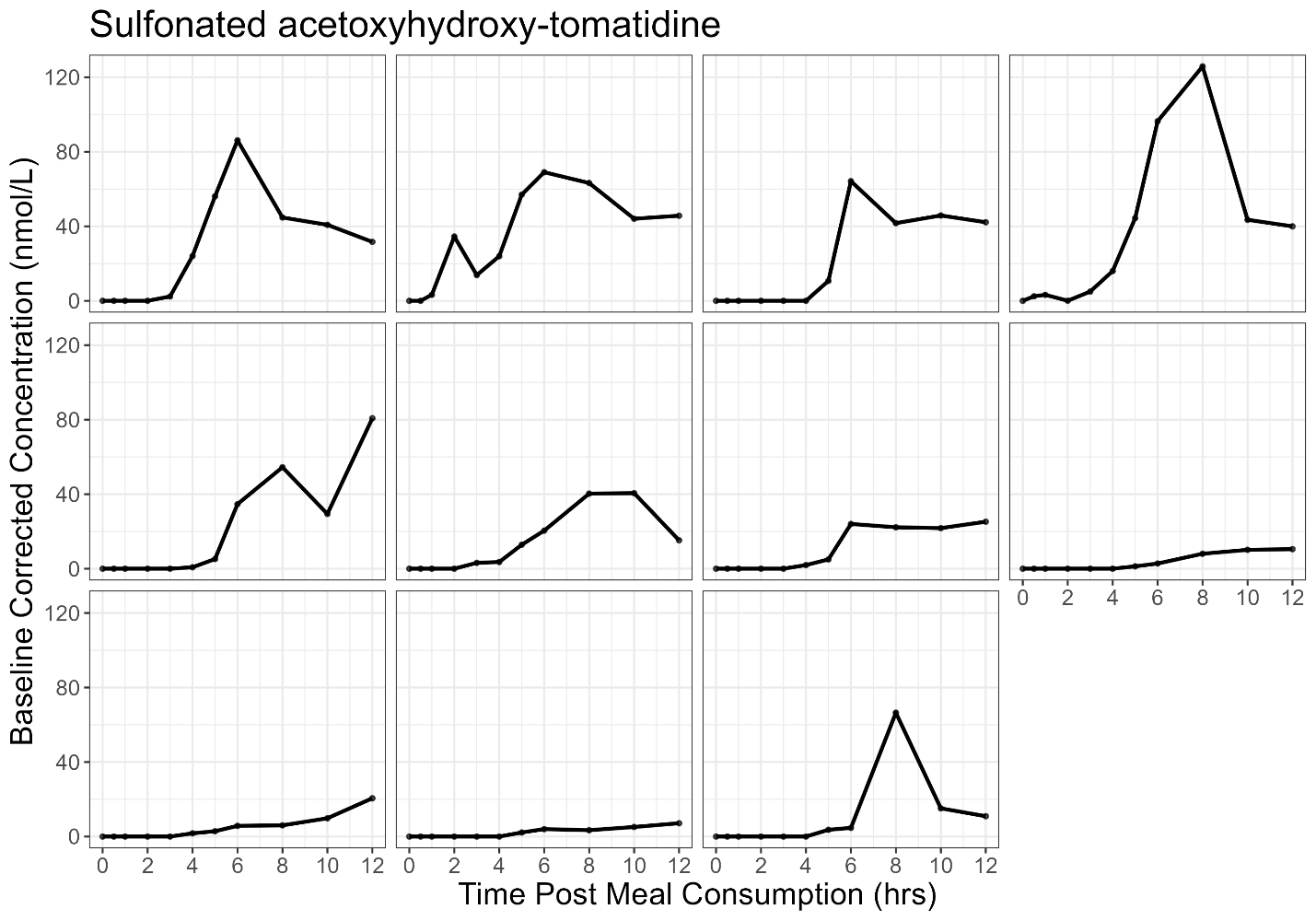

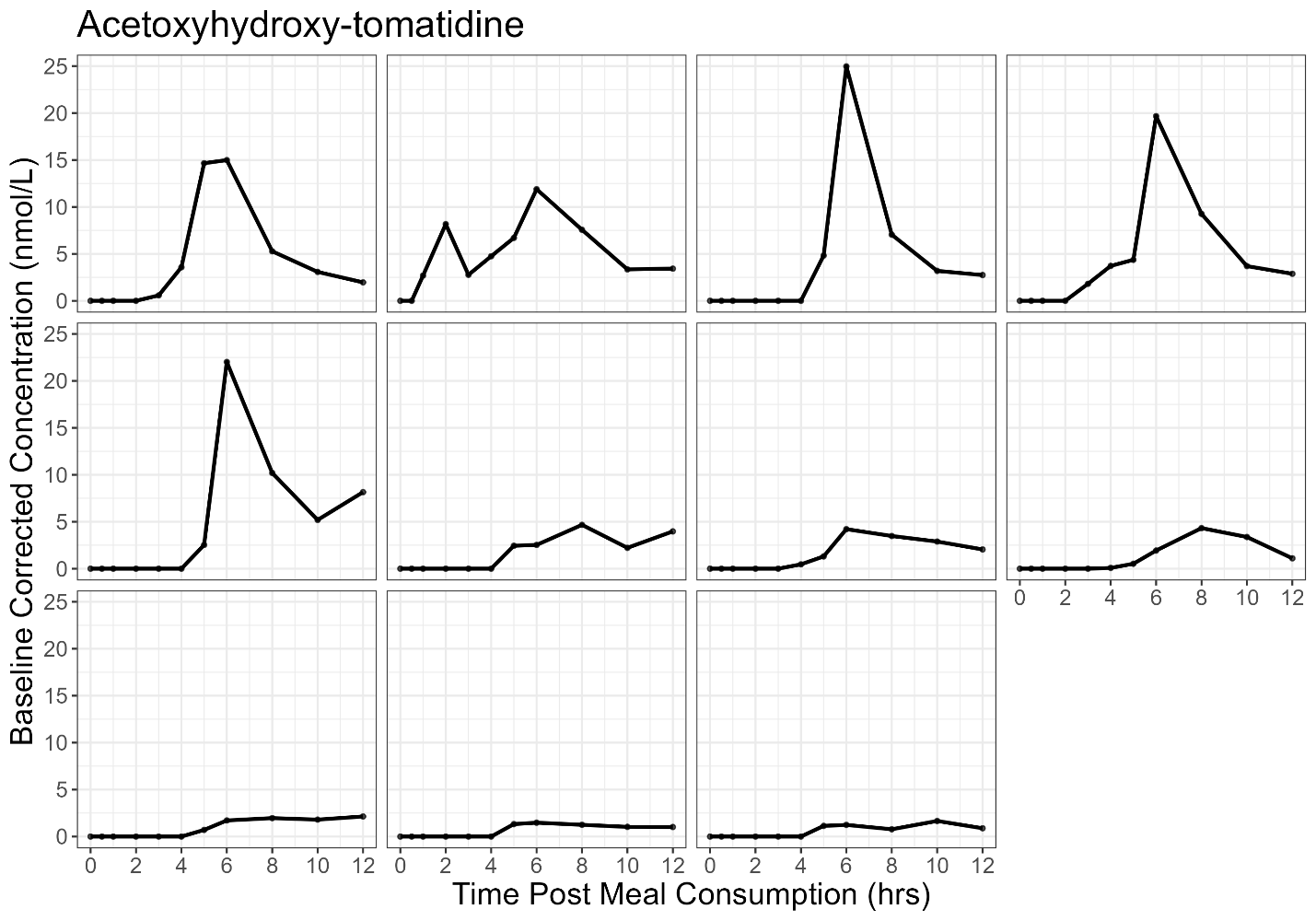

(E)

(F)

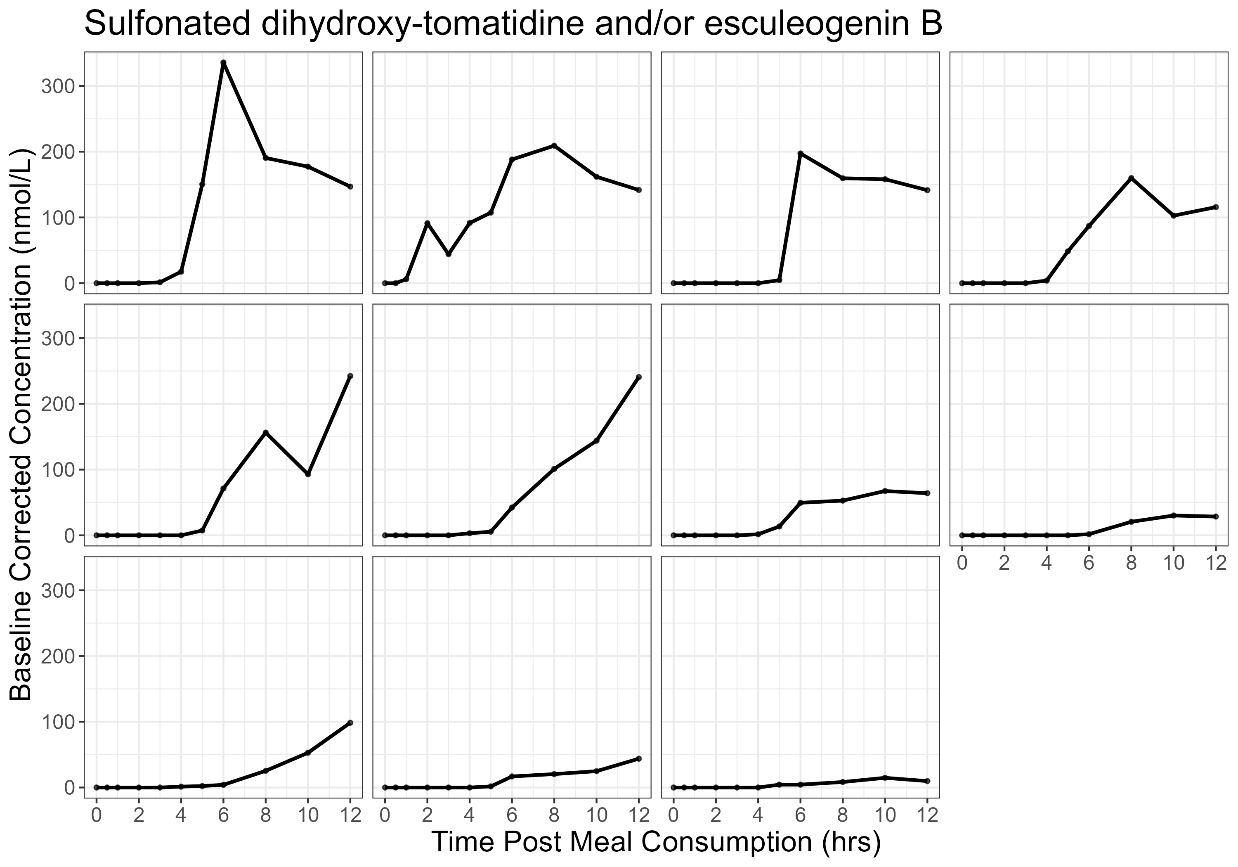

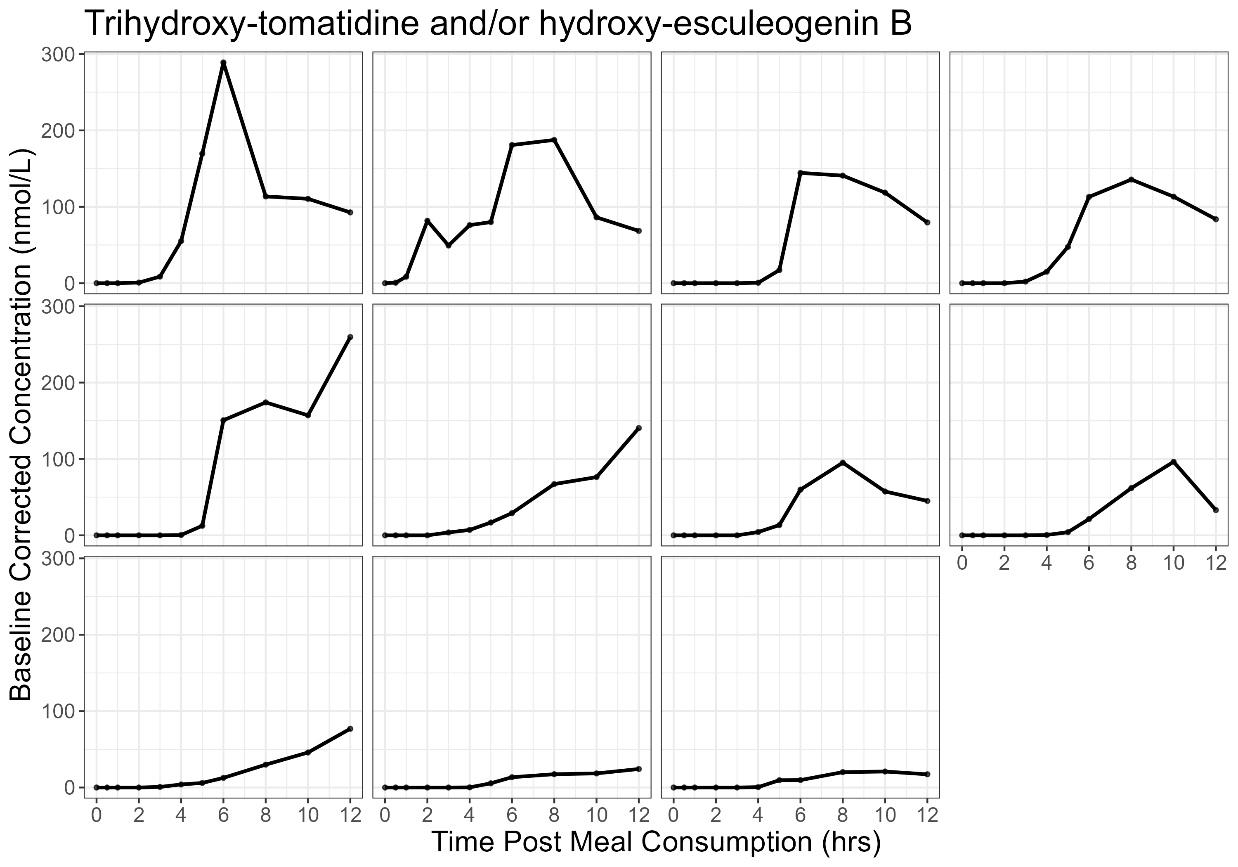

(G)

(H)

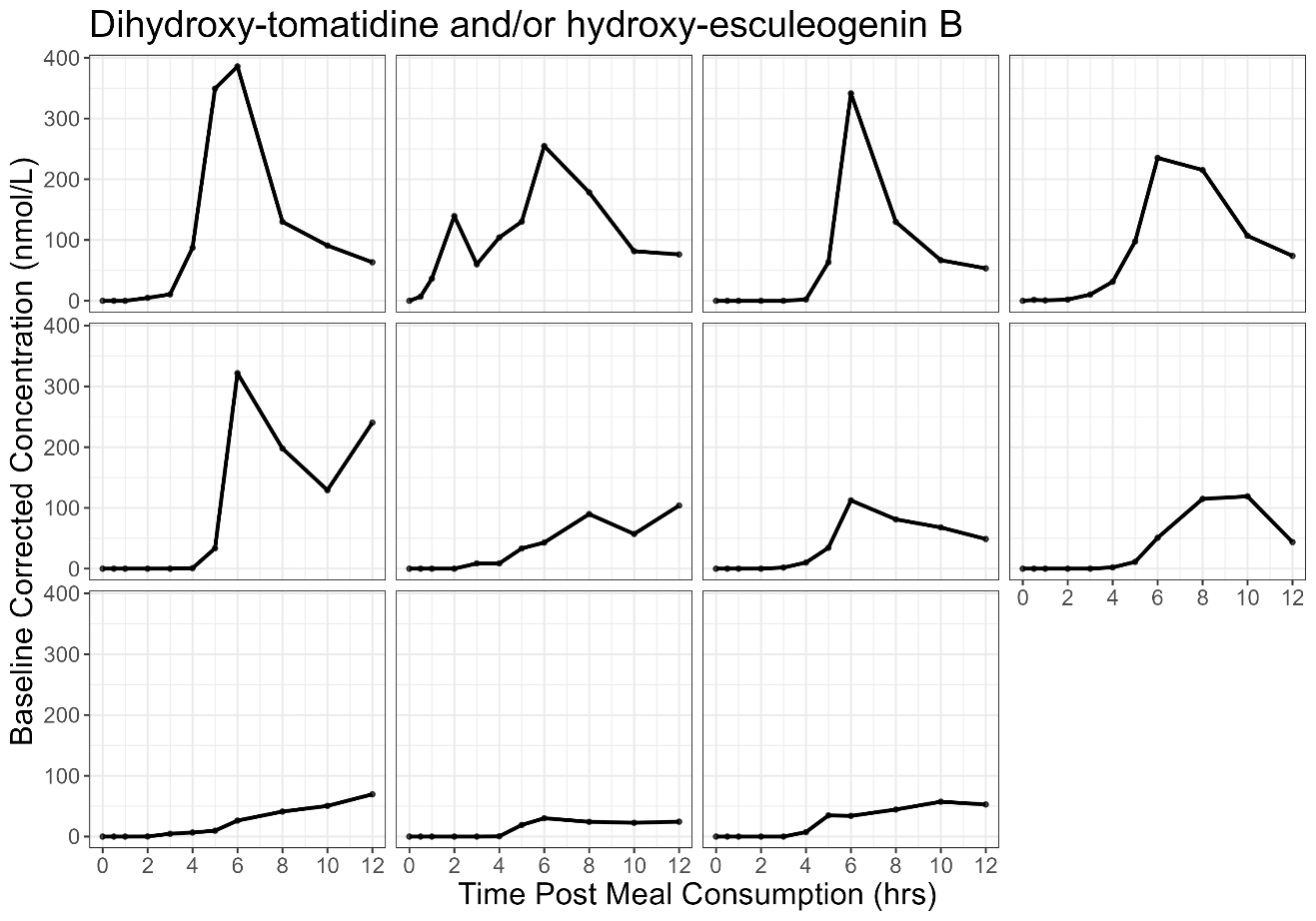

(I)
